# Barriers to sexual and reproductive health and rights of migrant and refugee youth: An exploratory socio-ecological qualitative analysis

**DOI:** 10.1101/2024.05.03.24306643

**Authors:** Michaels Aibangbee, Sowbhagya Micheal, Pranee Liamputtong, Rashmi Pithavadian, Syeda Zakia Hossain, Elias Mpofu, Tinashe Dune

**Author notes:** Corresponding author: Michaels Aibangbee.

## Abstract

**Purpose:** Migrants and refugee youths’ (MRY) sexual and reproductive health and rights (SRHR) is a global health issue. MRY tend to encounter adverse SRH experiences because of their limited access to and knowledge of SRHR services. Using a socioecological framework, this study examined the barriers affecting MRY’s SRHR.

**Methods:** A cross-sectional study utilising a participatory action research design was used. 87 MRY (ages 16-26, from 20 different cultural groups living in Greater Western Sydney, Australia) participated in the study and completed seventeen focus group discussions on MRY’s experiences of SRHR. Focus groups were co-facilitated by youth project liaisons for authenticity and validity. The data were analysed thematically and interpreted using socioecological theory.

**Results:** The findings identified socioecological barriers such as lack of awareness and access to services, sociocultural dissonance, and under-implementation of SRHR services. These barriers include cultural disconnects, language barriers, remote service locations, intergenerational cultural conflicts, and ineffective SRHR services. Key themes included traditional and institutional stigma, lack of SRH education, reliance on social media for SRH information and privacy concerns.

**Conclusion:** There is limited consideration of MRY’s SRHR and the impact of intergenerational discordance and stigma on MRY’s rights. The findings suggest the necessity for a collaborative SRHR strategy and policy design that empowers MRY’s agency across multicultural contexts.

## Introduction

Sexual and Reproductive Health and Rights (SRHR) plays a crucial role in the overall health and well-being of youth, particularly for those in migrant and refugee populations. A comprehensive understanding and provision of these rights are essential for a successful transition from adolescence to adulthood and positive long-term health outcomes for mental health, relationships and quality of life (Cheng et al., 2018; Iqbal et al., 2017). Yet research has shown that migrants and refugee youth (MRY) may not know where, how and when to access sexual and reproductive health (SRH) services or have the economic capacity to do so(Botfield et al., 2017). Studies indicate that MRY encounter barriers to contemplating and accessing SRH support and services which significantly impact their well-being. The barriers also extend to service providers who are not knowledgeable about the needs of MRY and, thus, lack the capacity to support them. For instance, Family Planning Services reported that of the low number of youth who accessed their services in 2012, 46% reported learning about them through word of mouth(Family Planning, 2016). This suggests that cultural stigma and misconceptions surrounding SRH may limit service accessibility, reinforcing reliance on informal networks for information about SRH(Family Planning, 2016). Further complicating these challenges are interpersonal factors such as language barriers, discrimination from non-migrant peers, and services lacking cultural awareness and safety. This contributes to an environment where MRY may feel uncomfortable discussing SRH issues (Atuyambe et al., 2015; Botfield et al., 2016).

Australia is one of the most multicultural countries in the world (Cavaleri et al., 2021). Therefore, it is important to examine migrant and refugee youths’ experiences of SRH in diverse areas of Australia. In the Australian context, Mpofu et al. (2014) argue that while SRH education is included in school curricula, it varies in breadth and depth across regions. Such educational inconsistency and social pressures exerted by peers, family, or the broader community may inhibit MRY from seeking SRH information and support (Mpofu et al., 2014). Moreover, existing educational resources are often developed without substantial input from young people, despite evidence suggesting that resources tailored to service users are more effective and accessible (Botfield et al., 2017; Botfield et al., 2016). This study explores these challenges further and propose strategies to improve MRY youth’s SRH service access by drawing on the experiences and perspectives of MRY.

Although SRHR is crucial to SRH and recognised as a key element of youth’s well-being, little is known about Australian MRY’s understanding of and experience with SRHR. This study seeks to contribute to the research gap by focusing on the perspectives of MRY and examining the role of these sociodemographic variables in the SRHR of MRY in Greater Western Sydney, one of the fastest-growing regions in Sydney with a migrant population of 50% (Fernandes et al., 2017; Haynes et al., 2021). No previous research has specifically mapped the socioecology of this demography with regard to SRHR. Such research findings can inform policies and practices to improve youth sexual and reproductive health wellbeing and, by extension, improve national health outcomes. Therefore, this is the first study that aims to answer the following research question: What are the socioecological barriers that impact MRY’s SRHR agency, decision-making, and wellbeing?

## Theoretical framework

### The socioecology of MRY’s sexual and reproductive health and rights

MRY face complex barriers in accessing, understanding, and implementing SRHR information. These barriers are interconnected at multiple levels, including individual, interpersonal, institutional, and societal. Bronfenbrenner’s socioecological theory (1979) provides a practical framework (Fig 1) for understanding these barriers as it considers the five interconnected layers of environments surrounding the individual: the individual, microsystem (interpersonal), mesosystem (relationships between microsystems), exosystem (institutional), and macrosystem (societal) (Bronfenbrenner, 1979).

**Figure 1.**
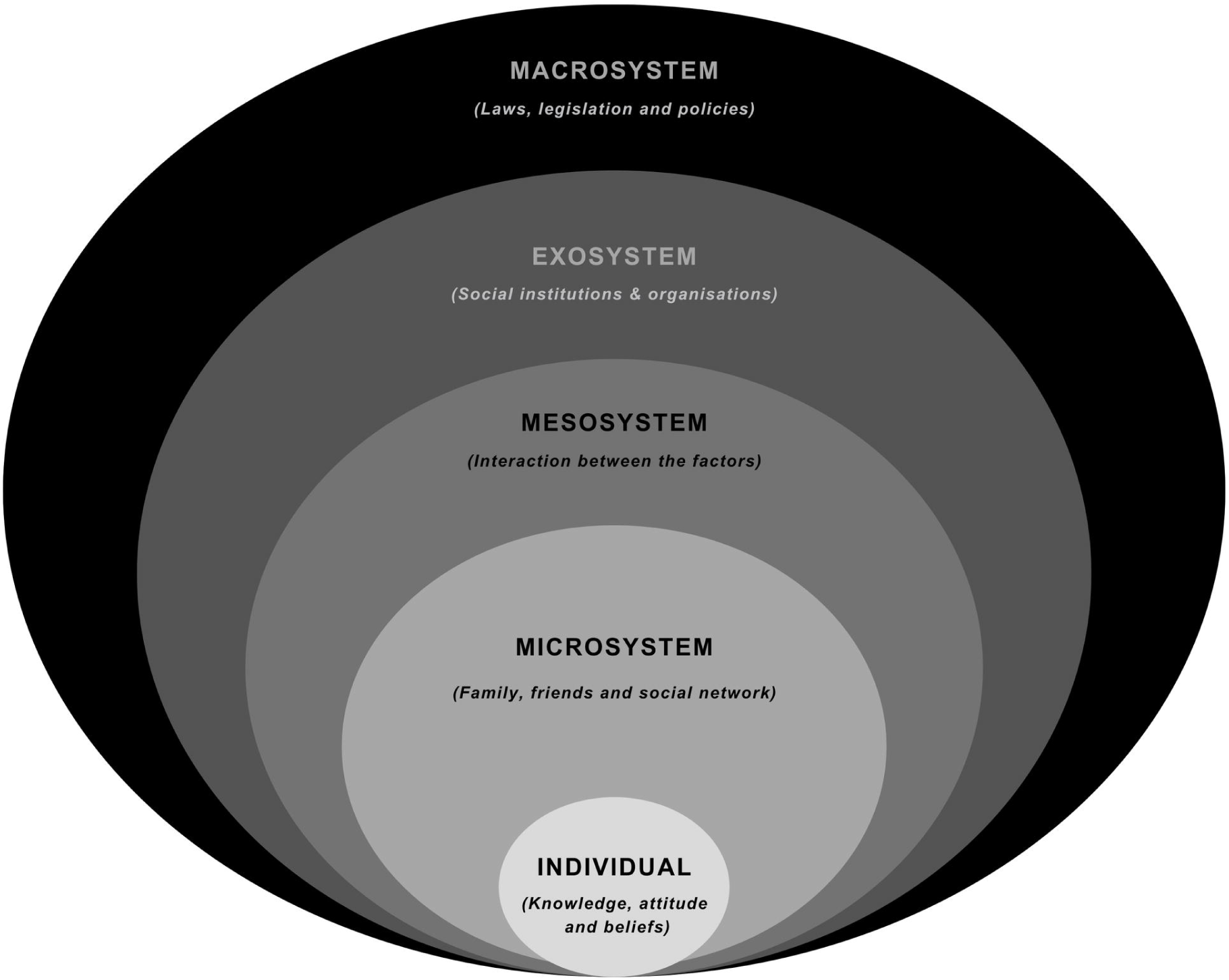
Bronfrenbrenner’s Socioecological framework (1977)

Factors such as age, gender, sexual orientation, education, personality, length of time in the host country, and acculturation strategy can affect MRY’s SRHR access and implementation (Ganczak et al., 2017; Tirado et al., 2020; Yakushko et al., 2008). These factors, along with socioeconomic status, language barriers, and traumatic experiences, can influence the interpersonal, organisational, and societal expectations placed on MRY, which affects their engagement with SRHR (Dune et al., 2017; Metusela et al., 2017; Rawson & Liamputtong, 2010; Ussher et al., 2017).

From the mesosystem and exosystem perspectives, MRY may have limited knowledge of what, where, how, and when to access SRHR services (Botfield et al., 2017; Dune et al., 2017; Ivanova et al., 2018; Mulubwa et al., 2020). Botfield et al.’s (2016 and 2017) studies show that SRH education provided to young people is often developed without input from the demography, which may result in limited accessibility and utilisation of services.

At the macrosystem level, MRY navigate cultural influences from their culture of origin and the host country, leading to intergenerational conflicts, fear, and structural social misunderstandings around SRHR (Berry, 2005; Dey & Sitharthan, 2017; Kagitcibasi, 2007; Phinney, 1990). This can result in disengagement from SRHR and negative physical and psychological consequences such as exposure to sexually transmitted infections, unplanned pregnancies and poor mental health (Habtamu & Adamu, 2013; Herd et al., 2016). Institutional and societal contexts also impact MRY’s ability to access SRHR information and services. According to research, in Australia, migrant and refugee populations often experience SRHR services as culturally inappropriate or insensitive, leading to limited engagement and poor outcomes (Hawkey et al., 2021; Heslehurst et al., 2018; Metusela et al., 2017; Napier-Raman et al., 2023). Factors such as limited access to interpreters, a lack of continuity of care, and perceptions of unhelpful or uncaring staff contribute to this problem (Botfield et al., 2017; Mengesha et al., 2017; Riza et al., 2020).

When categorised using the socioecological model, the factors that emerge at the microsystem level include immediate family, school, and friends, while the mesosystem level examines the interactions between these factors (Napier-Raman et al., 2023). At the exosystem level, healthcare systems, extended family, and government policies play a role in shaping MRY’s approach to SRHR, while the macrosystem level encompasses attitudes, ideologies, culture, and religion, all of which influence MRY’s reproductive health decisions in Australia (Napier-Raman et al., 2023). Mpofu (2018) and Amroussia et al. (2022) agree on the significant impact religion can have on MRY’s SRHR beliefs and behaviours (Amroussia, 2022; Mpofu, 2018). Cultural concentration, in which migrants maintain a connection to their culture while adapting to the host country, can bring both opportunities and challenges for MRY in renegotiating relationships and family structures in a new cultural context (Dune et al., 2017; Rawson & Liamputtong, 2010; Saleem et al., 2017; Villa-Torres & Svanemyr, 2015). Therefore, by exploring these various factors within the socioecological framework, we can comprehensively understand the complex interplay of factors that influence the MRY’s approach to SRHR.

## Methods

### Study design

This paper reports on the findings from qualitative data collection with MRYs aimed at capturing the depth, nuances and complexities of MRY’s SRHR experiences and perspectives, which are often lost in quantitative approaches. The findings presented in this paper are from a larger mixed methods study funded by the Australian Research Council (DP200103716) entitled *Migrant and Refugee Youths Sexual and Reproductive Health and Rights*, which sought to examine MRY’s access, decision-making and utilisation of SRHR services and to develop a model for service implementation. The broader study used a Participatory Action Research (PAR) design to engage and empower MRY, enabling them to identify research questions, participate in data collection, and provide useful information to facilitate change. PAR is a research methodology that prioritises the active involvement and collaboration of communities in research and has been applied in health research to address complex social problems that require transformation (Liamputtong, 2006; Reason & Bradbury, 2008). Applying the PAR methodology in SRHR research ensures that researchers, youth project liaisons (YPL), advisory committee members (ACM), and MRY come together as partners to define issues, co-design solutions, and implement changes. Liamputtong (2020) notes that this approach is critical for sensitive SRHR issues requiring confidentiality and respect for diversity (Liamputtong, 2020).

## Recruitment and sample

The recruitment process for this study was executed in multiple stages to involve the youth participants in developing the research. Recruitment commenced on 01 June 2020 and ended on 12 June 2021. Three groups of participants (ACM, YPL, and MRY) were drawn from a diverse range of racial, ethnic, religious, socioeconomic, educational, sexual, and geographical backgrounds. A total of 87 young people participated in the study (YPL, *n=8*; MRY, *n=79*). Demographic information was obtained from 75 youth participants, with 56 (65.12%) identifying as females and 19 (22.09%) as males. The youth participants were between 15 and 29 years old (Appendix A shows the detailed demographic profile).

### Advisory Committee Members (ACM)

ACM includes individuals from key stakeholder groups (such as community managed organisations, community leaders, influencers and workers) and health professionals with pertinent careers and experiences. Their contributions were integral to implementing the PAR framework in this project. They also assisted with convenience sampling and recruiting YPLs who had active involvement with the migrant and refugee communities in Greater Western Sydney.

### Youth Project Liaisons (YPL)

The YPLs served as relatable liaising peers to other MRY who participated in the study. The function of YPL also helped to mitigate any perceived power differential between the researcher and the youth, given that the YPL were in the same age demography as the MRY. Hence, YPL taking an active part in the PAR process helped mitigate their perception of the researcher as an outsider.

### Migrant Refugee Youth (MRY)

As the focus of this study, MRY offered invaluable qualitative data on their experiences and understanding of SRHR, which was necessary for this project to meet its aims. Subject to the inclusion criteria (Table 1), MRY were recruited via a range of sources including ACMs, YPLs, social media advertisements (Facebook, Instagram, and Snapchat), community organisation newsletters, printed materials posted at Western Sydney University campuses, and notice boards in shopping malls, churches, and community organisations in Western Sydney. A diverse sample of young people were recruited across racial, ethnic, religious, socioeconomic, educational, sexual, and location groups.

**Table 1.** Focus Group Description and Inclusion Criteria.

| YPL-Led SRHR Focus Group |  |
| --- | --- |
| Focus Group Description | The focus groups (average of 60 mins per session) explored the youths’ understandings, experiences, barriers and facilitators relating to SRHR. |
| Inclusion Criteria | The inclusion criteria for youths in all parts of the study are:<br>1) being aged 15 to 26,<br>2) self-identifying as a migrant or refugee and<br>3) living in Greater Western Sydney for the past 12 months. |

### Data collection

Seventeen focus groups were co-facilitated by the first author, ranging from 60 to 90 minutes, with an average time of 60 minutes. The focus groups were held during the COVID-19 pandemic, which resulted in focus groups being convened mostly online via Zoom® between November 2020 and June 2021 with participants located in their preferred spaces. Three of the focus group sessions were conducted face-to-face, two of which were held simultaneously in two different spaces at a local community-managed organisation in Greater Western Sydney. The sessions were audio-recorded and professionally transcribed verbatim using Trint® software, followed by an extended manuscript verification by the research team.

### Procedures

Participants were recruited, organised, and facilitated through YPLs, who recruited their peers via convenience sampling, and from the research team’s networks (17 FGs, ranging from 2 to 10 participants per focus group, *n=*79 youth). The focus group questions (Appendix B) were developed in collaboration with ACMs, YPLs, and the research team. The focus groups explored MRY’s understandings, experiences, barriers, and facilitators related to SRHR.

Central to the PAR methodology, YPLs engaged with the project at various key phases and were trained to recruit and engage MRY and co-facilitate focus groups at the data collection stage. The YPLs also participated in an annual focus group (60 minutes) over three years to gain insight into their experiences engaging in a PAR project and their perspectives on the project’s alignment with human rights principles. The annual focus groups were audio-recorded, transcribed, and analysed using the existing data analysis protocol.

### Ethical considerations

The Western Sydney University Human Research and Ethics Committee approved the ethics framework (H13798) before the study commenced. The initial plan was to conduct the focus group through in-person sessions; however, due to COVID-19 restrictions, the Zoom® platform was employed as an alternative.

### Consent

A participant information sheet (PIS), consent information, and inclusion criteria were distributed to the participating MRY to ensure their understanding and consent before participating in the study. Completed consent forms and consent by assent were also accepted for convenience and to minimise paperwork, particularly for some disadvantaged youth. At the start of each workshop or group session, the facilitator sought additional verbal consent to confirm participants’ willingness to participate in the focus group.

### Data Analysis

The data from the focus group sessions were thematically analysed (Braun & Clarke, 2019) by the first, second, and last authors. Thematic analysis was performed by identifying topics and substantive categories within the participants’ accounts in relation to the study’s objectives. Pseudonyms were used to protect participants confidentiality. Quirkos® is an intuitive qualitative data management software that assists researchers in coding and analysing qualitative data (Quirkos, 2021). Quirkos® was used to ascertain topical responses and emergent substantive categories, coding for word repetition, direct and emotional statements, and discourse markers, including intensifiers, connectives, and evaluative clauses (Braun & Clarke, 2019; Liamputtong, 2020). YPLs also attended a workshop where they were taught basic qualitative analysis principles and then worked in groups of two to analyse two of the 17 focus group transcripts. The resulting codes were added to the thematic analysis of the qualitative data.

## Results

This study examined the various socioecological barriers (see Fig 2) that impact MRY’s SRHR using Bronfenbrenner’s socioecological systems framework. A key focus was understanding how MRY navigated their SRHR against the identified factors. Thematic analysis of the collected data revealed themes that provided insight into MRY’s understanding and experiences of SRHR in line with the following socioecological levels: Micro-, Meso-, Exo- and Macrosystem levels.

**Figure 2.**
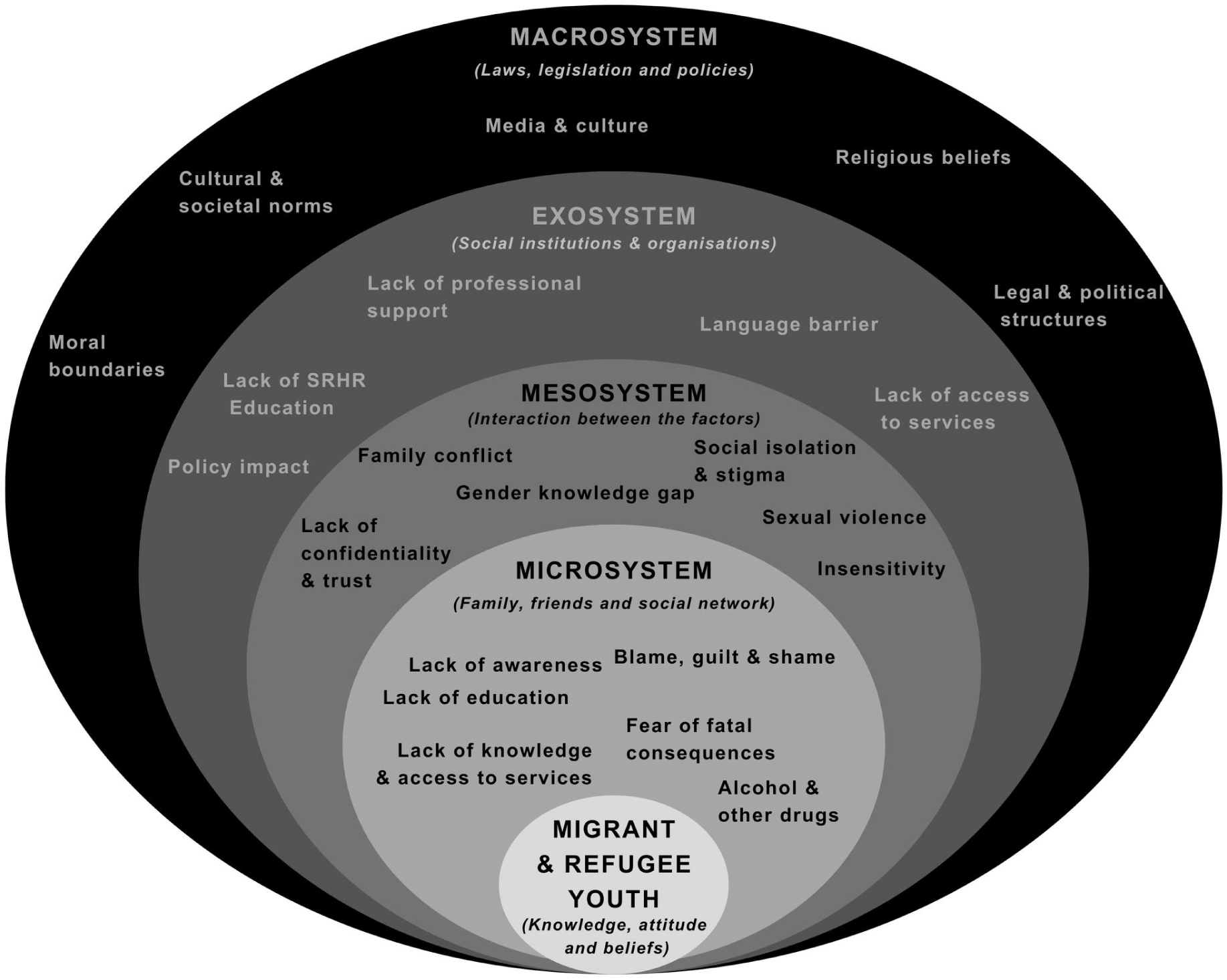
Socioecological barriers affecting MRY’s SRHR using Bronfenbrenner socioecological framework (Sydney, July 2023)

### Microsystem level

Various barriers affecting MRY’s SRHR were identified at the microsystem level including lack of awareness, lack of education, fear, blame, guilt, and shame, and lack of knowledge and access to services. These barriers are often exacerbated by challenges of their migratory status and play a crucial role in how MRY navigate their SRHR. These factors emphasise the complexity of SRHR issues at the individual level, indicating how different personal experiences, feelings, and the context of education and awareness can shape an individual’s understanding and behaviour regarding SRHR.

#### Lack of awareness and access to services

MRY often come from cultural backgrounds where open discussions about SRHR are discouraged or non-existent (Thornicroft et al., 2008). Some participants reflected a lack of awareness regarding SRHR, particularly due to cultural or societal norms. For instance, one participant noted, *"I come from a country where… you weren’t taught about reproductive health a lot… there were young, 16, 17-year-olds who didn’t know that sex led to falling pregnant…" (Sarah, F, Sri Lankan)* revealing how a lack of awareness can lead to basic misconceptions about reproduction. Such a lack of awareness may contribute to risky sexual behaviour and prevent MRY from seeking the necessary reproductive health services. Absence of knowledge was also seen in the understanding of concepts such as consent, with another participant stating*, "A lot of people don’t really know what consent is still… maybe it’s just where they’re from or how they were raised that they don’t truly grasp the concept of consent" (Lupita, F, Fijian)*.

Furthermore, participants expressed concerns about not only the lack of knowledge about SRHR but also the lack of access to services. One participant acknowledged, *"Just the lack of knowledge [about services] is what stops me… possibly not me, but maybe incorrect knowledge… that could be something that would stop someone from being able to protect their sexual reproductive health" (Loretta, F, Indian).* The participants’ experiences underscored the impact of this lack of knowledge, unfamiliarity with the healthcare system, and access to the individual’s ability to protect their sexual and reproductive health. A participant expressed that*, “it would definitely be helpful if these [SRH] centres were located closer or more around the western Sydney area, and they’re more easily accessible if they had proper funding” (Johnny, M, Indian)*.

#### Lack of SRHR education

Lack of adequate SRHR education was a recurring theme in the data across all the focus group sessions. Participants unanimously reflected on the ineffectiveness of SRHR education in various schools they attended. One participant stated, *"There’s a disparity between how much about sexual health and consent and stuff is taught in high school compared to… my friends from other areas in Sydney" (Jas, F, Indian).* Participants emphasised a disparity in the amount of sexual health education received in school, adding*, “Some of my friends [like] didn’t even know [like] stuff about consent until we were in like year 11 or year 12, and we had to [like] find that ourselves. [Like], no one really sits you down and explains to you [what] anything about sexual health. [Like] in PE, it was [like] a topic on it (SRH) or a couple [of] topics on it, but they don’t spend a lot of time on it" (Ethan, M, Chinese).* The lack of school-led instruction about SRHR resulted in individuals feeling unprepared and unsafe, as indicated by another participant, *"I don’t think it’s appropriate for anyone under 18 to have sex when they don’t when like, school doesn’t really even teach you how to do it safely" (Jas, F, Indian).* This implies that a lack of education can leave MRY ill-equipped to make safe and informed decisions about sexual health.

#### Fear of fatal consequences

Fear, often stemming from inadequate or fear-based sexual education, can have detrimental effects on MRY’s approach to SRHR. Fear of the consequences of sexual activity was evident in participants’ accounts. They emphasised being taught to avoid sex due to the risk of contracting diseases, such as HIV, rather than being educated on safe practices. One participant put it quite starkly: *"Yeah, the point of the AIDS presentation is to, ‘don’t have sex because you will get AIDS… just don’t have sex because you will die’" (Angelene, F, South Indian)*.

#### Blame, guilt, and shame

MRY highlighted the potential culturally or socially instigated feelings of shame, guilt, and blame regarding SRHR, making it difficult to access necessary services or communicate these issues (Baigry et al., 2023; James et al., 2020). This was mirrored in the participants’ reflections, where societal judgement, internalised shame, and feelings of personal blame hindered their willingness to seek help. For instance, participants mentioned, *"Like, you know, women not going to get these checkups …marital rape, you know this could be happening. But they’re like, ’I don’t want to bring this up’ because [like] the women end up being shamed’" (Lupita, F, Fijian).* Another participant stated, *“I’d prefer to distance myself from someone from [like] a similar cultural background just because I think I’d always feel somewhat like judged or I’d always feel like the taboo of, like my cultural upbringing in the commentary of that doctor” (Zantla, F, Bangladesh).* Another highlighted the blame-laden guilt where the MRY is expected to know better, stating, *“I have the knowledge I learned about it in school, but… it’s always preventative but it’s never kind of like, ‘OK, now, say you do have it (STI), what are your options? How do you go about this?’ It’s always preventative and therefore, if you get it, it almost implies that it’s your fault ‘…that’s your responsibility, that’s on you’” (Angelene, F, South Indian)*

#### Alcohol and Other Drugs (AOD)

Participants highlighted that using alcohol and other drugs impacted decision-making regarding SRHR. One participant described how being under the influence of alcohol makes it harder to think clearly, stating, *"When I drink, it’s like you, I don’t think of everything. You’re kind of just in the moment, so you forget certain things, " (Moana, F, Fijian)* thus increasing the likelihood of unsafe sexual behaviour, resulting in unplanned pregnancy, psychological harm and STI. Another participant highlighted the effect of AOD on the reproductive system, stating, *“If you took drugs, alcohol or certain things, they can affect your reproductive organs, which is a very complicated system” (Evelyn, F, Indian)*.

### Mesosystem level

At the mesosystem level, interactions between various microsystems, such as family and school, peer groups, and healthcare professionals, greatly impact the SRHR of MRY (Coatsworth et al., 2002; Neal & Neal, 2013). The various factors indicated by the data are as follows.

#### Family conflict

Conflicts within families often arise because of generational and cultural differences in sexual health. Family conflict was found to create a barrier to open discussions on sexual and reproductive health. A participant noted, *“Even though I’m growing up, they may still see me as a child so they don’t really - it’s hard for them to catch up, really, in their mind like, ‘oh its time, and she’s old enough, and she’s mature enough’" (Leticia, F, Nigeria).* As the family is often the primary source of information about health and personal development for young people, conflict can prevent MRY from accessing critical information, leading to misunderstandings and potentially harmful misconceptions about SRHR. For example, one participant stated, *“If, maybe if your parents found out you were researching about sexual health… they’re like’, what’s going on?’” (Leticia, F, Nigeria)*. While another added, *“Yeah, even though I’m* [legally an adult]*, yeah, they’ll still punish me, because in my family, they still look at me as a little child” (Moana, F, Fijian).* This illustrates how parents often struggle to reconcile their child’s evolving maturity and independence, which can hinder open dialogue about SRHR within the family. In addition, the absence of support from family members can pose a significant challenge in managing SRHR. One participant described her struggle to maintain secrecy about using contraceptives at home: *"I had to keep everything very secretive if I was using a pill, if I was using contraception" (Angelene, F, South Indian).* Without support from their families, MRY may face difficulties in accessing SRHR services or in understanding their rights. Further, *“I always felt like, protecting my health or maintaining my health in a household […] that never really talks about it, is me being in trouble. It’s just weird sort of like association, where I went out of my way to make sure I was safe or I’m doing the best for my body, but if I get caught, I’m in trouble” (Angelene, F, South Indian).* The secrecy due to the lack of family support may further contribute to unsafe SRH behaviours and poor health outcomes.

#### Social isolation and stigma

The data showed that stigma around sexual experiences can exacerbate social isolation, limiting access to peer education and support around sexual health issues which means fewer opportunities for MRY to share and learn from their peer experiences. A participant noted, *“There’s a lot of social stigmas around sex in general, [like] there’s a stigma around having a lot of sexual partners, there’s a stigma if you haven’t had any” (Marley, F, Chinese).* Reflecting on their experiences, a participant talked about another student who was *“… really socially awkward or isolated or [like] just feel alone or feel like they can’t trust anyone; I think it would be an even more challenge for those kids” (Zainab, F, Pakistani).* This lack of interaction can restrict MRY’s understanding of SRHR, which may lead to unsafe SRH practices and negative health outcomes.

#### Gender knowledge gap

This refers to the disparity in understanding and knowledge of sexual health between genders, which could lead to misconceptions about SRHR, potentially contributing to unsafe practices and reinforcing gender inequalities. The gap can be widened by the influence of pornography and the media, which often portray unrealistic and harmful views of sexual health and relationships. Referring to the masculine gender, one participant emphasised the perceptual limitation, *“I’m so surprised, I’m like, ‘by your old age, like 20, 30 years old, how do you not know? I can’t start my period on command.’ Or they think your vagina is supposed to smell like flowers.” (Dipthi, F, Pakistani)*. Another participant added, *“And that really bothers me because a lot of young girls get like insecure, and dirty men with their fingers, they’re touching it. And I’m just saying, like, you can get a yeast infection. It’s not okay. Like they think you’re supposed to smell pretty, but they don’t do the same thing about their situation down there. Like it has to be both ways; [participating]in sex health classes where they show that it’s not a bad thing, like you’re supposed to smell like that. It’s a vagina. It’s moist down there. (Monique, F, Bosnian).* Another participant expressed a slightly different take, *“I think taboo regarding sexual health that exists within cultural communities is, in my opinion, I think it’s a result of misogyny most of the time… I think sexual health is targeted towards women more than men” (Darlene, F, Ghanaian)*.

### Sexual violence

Experiences or exposure to sexual violence can severely impact SRHR, often traumatising and influencing future sexual behaviour and health. As one participant stated, *"Like if you have to pester someone, where you have to like try and push them or influence them into doing something that they’re not willing. For example, like even if you’re like under the influence of alcohol and you and the other party can’t consent because they’re not in like a sober state of mind, like that’s still coercion, you know, like you’re forcing someone to do something that they don’t want.” (Moana, F, Fijian).* In agreement, other participants iterated, *“Yeah, that can put a lot of pressure on you, like mentally if you haven’t consented to something” (Rita, F, Liberia).* And another, *“They make you think you want to. Yeah, they asked repeatedly. They don’t drop it. They keep asking and asking, and eventually you say ‘yes’ because you’re tired of saying ‘no’ because saying no isn’t working. So you say ‘yes’ and then give in. So that isn’t consent.” (Natalie, F).* Exposure to sexual violence can have devastating effects on the SRHR of MRY, increasing the risks for STIs, unwanted pregnancy, and long-term psychological harm. Another instance of sexual violence was highlighted around honest conversations about the use of protection or contraception. A participant reported, *“I heard about [like] how some guys [like] they would say, ‘yes, [like] I’m using the condom,’ and then even the girl would say, ‘OK, you’re using it,’ and then they would just not use it; [like] they would manipulate the other partner” (Aileen, F, Pakistani)*.

### Lack of confidentiality and trust

At the Mesosystem level, the lack of confidentiality and trust significantly influences MRY’s experiences and perceptions of SRHR. This lack of trusted guidance can foster feelings of isolation and anxiety, as illustrated by one participant who admitted, *"I was just like, paranoid about it at one point, and then obviously I couldn’t talk to my parents and that’s about it” (Johnny, M, Indian).* Another participant expressed frustration over the scarcity of reliable resources or authoritative figures to guide them through the complex maze of SRHR, stating, *"There’s no sort of representation of how to do that…there’s really nothing you can -, and your own friends or people around in your school and we’re all trying to navigate this, but there’s no sort of guiding person who can talk to you about these sorts of things” (Leila, F, Lebanese)*.

Additionally, the intersection of personal and cultural spaces can further complicate these experiences, causing MRY to avoid seeking help because of cultural stigma or fear of judgement. A poignant example of this came from a participant who recounted their uncomfortable experience with a family-acquainted doctor. She stated, *“I was going to a doctor, and this was so awkward. As soon as I stepped into that room, she asked me, ‘oh, how’s your family? and all of that.’ That just alienated me from the whole medical purpose that I wanted to come in there for because I don’t want that kind of, you know, I don’t want a family friend or anyone from my specific communal ethnic group knowing about my medical concerns and needs” (Zantla, F, Bangladeshi).* These experiences underline the critical need for increased representation and confidential services in SRHR, specifically catering to MRYs.

#### Insensitivity

Insensitivity towards SRHR, especially from educators, service providers, or familial adults, can further perpetuate the stigma and fear associated with SRH, discouraging MRY’s help-seeking behaviours. Insensitivity can lead to feelings of shame and guilt, furthering the harm caused by lack of education and support. Reflecting on a personal experience, a participant recalled, *"They don’t even acknowledge that it’s like a personal and sensitive topic; …you don’t really get that one on one contact or even just make it feel like a personal experience" (Rollah, F, Lebanese).* Another participant added, *“I think it’s upsetting because it’s an upsetting situation,,, and then you’re just left to be like, ‘you know, it is what it is’” (Ayelen, F, Nigeria).* This lack of sensitivity can deter MRY from seeking necessary information and support for their SRH.

#### Exosystem level

At the exosystem level, the environment surrounding MRY often indirectly impacts their attitude towards SRHR through several broader societal factors, including the absence of professional support, language barriers, policy impacts, inadequate SRHR education in the school curriculum, and lack of access to services (Botfield et al., 2017; Chattu & Yaya, 2020; Dune et al., 2017; Malia et al., 2021). The following themes emerged considering the exosystem level.

#### Lack of professional support

This is a cross between the meso-and exosystem in the lack of professional support, which often means that MRY lack the necessary guidance in navigating SRHR services. This hinders the MRY in making informed decisions about SRH. A male participant reflected on a similar situation, stating, *“There’s no sort of representation of how to do that, like, I can’t really think of, other than pornography, there’s really nothing you can [do], and your own friends or people around in your school, and we’re all trying to navigate this, but there’s no sort of guiding person who can talk to you about these sort of things” (Lee, M, Chinese).* Rollah (F, Lebanese) added*, "No one really has the follow-up conversation. I think that’s a massive gap in sexual health and reproductive health, education.”*

#### Language barriers

Language barriers further complicate the situation, limiting the MRY’s ability to understand and engage with SRHR services in their new home countries. Language barriers affect accessibility and understanding of SRHR (Endler et al., 2021; Tirado et al., 2020). MRY often find it challenging to explain complex concepts like consent or contraception when delivered in a language they are not familiar with. Nasrat *(M, Indian)* stated, *"Things that are easy for us to talk about in English, you can’t explain that to like maybe older people or recent migrants or refugees,"* pointing out that discussing SRH topics is already challenging and more challenging when a third party is required to interpret it or when one is required to find the right words to express their presentation in a different language.

#### Policy impact

Policies that are not designed with the unique needs of MRY in mind can contribute to these challenges. An example is the taxation of menstrual health products, disproportionately affecting financially disadvantaged MRY (Tirado et al., 2020). Some MRY acknowledged the challenge, summed up by another stating, *"… not everyone has access to, you know, menstrual hygiene products or anything like that" (Sila, F, Lebanese).* This implies that policies do not adequately address the needs of the MRY in terms of the SRHR. Another participant stressed the difficulty in navigating settlement as a new arrival, combined with SRHR challenges and its existing perspectives, complicated by the trauma of the migratory process. She stated, *“if you come from another country, there’s a different culture there. …When your parents are from somewhere else and then migrated to a new place, it’s a bit full on” (Jael, F)*.

#### Lack of SRHR education in the curriculum

The lack of comprehensive SRHR education in the school curriculum, which typically does not include direct input from young people or MRY, leads to misunderstandings and misinformation about MRY’s SRHR. The lack of SRHR education in the curriculum has led to misconceptions and ignorance. MRY commented, *“You shouldn’t get hurt as a kid. You should get taught this information because it is important to your health. You don’t get taught” (Darlene, F, Ghanian).* Another added*, "I think a lot of young people, like, unfortunately, because of the lack of education, they sometimes, um, detriment the sexual reproductive health without even realising it” (Jas, F, Indian),* and *“Teaching abstinence only is unsafe” (Monique, F, Bosnian).* These statements imply the need for an overhaul of the educational curriculum with SRHR content that is beneficial to MRY taking their needs into consideration.

#### Lack of access to services

MRY often find it difficult to access SRHR services due to various factors, including limited knowledge about service availability, stigma, and logistical issues, such as scheduling conflicts with school. For instance, one participant noted, “*Around access to clinics, [like] when I think about when I was first exploring, [kind of] sexual relationships, it was in high school, and it was a lot of pressure, and I didn’t have access; [like] you’re at school for most of your week and then…, you have a curfew, or you have families [like] you get picked up from school. So there’s never really a chance to go to a clinic. I didn’t have access to go out of my current routine because my parents or family weren’t supportive, that wasn’t a thing. I can’t skip school, obviously. So that was never really accessible to me in high school” (Angelene, F, South Indian).* The data also show that the relative invisibility of SRH services, such as abortion clinics, further amplifies SRHR barriers. As one MRY noted, *"No one really [like] advertises about it. [Like] you never see it when you walk down the street. You see [like] Cancer Council clinics [like] you can get your skin checked. Those clinics have labels, right? Have you ever seen [like] one for abortion?" (Marley, F, Chinese)*.

#### Macrosystem level

At the macrosystem level, numerous cultural factors often intersect to influence the SRHR of MRY. Cultural norms and beliefs from both their country of origin and Australia shaped their experiences. This dual influence can result in internal conflicts, especially if the two cultures have different attitudes towards SRHR. In many cases, the cultural norms of their country of origin, which often discourage discussions about SRHR, may conflict with the more open attitudes seen in the Australian culture (Botfield et al., 2016; Dune et al., 2017; Rawson & Liamputtong, 2010).

#### Cultural and Societal Norms

The data shows that cultural beliefs and societal norms can sometimes foster an environment where discussions on sex and reproductive health are stigmatised, consequently limiting young people’s access to essential information and resources. For example, an MRY expressed, *"I remember, as a kid, whenever we watched TV, and anyone was kissing, and your parents are there, you have to act like you’re embarrassed like you hate [it], and it’s disgusting" (Moana, F, Fijian).* Another participant summarised the pervasive influence of these norms, stating,

*“I think, in the cultural perspective… if you’re raised, um, surrounded by people who are the same culture as you, you don’t need to question anything, you just go with the flow and you go with the social status because everyone around you has the same culture, the same upbringing so you don’t question anything outside of that little box…” (Darlene, F, Ghanian).* In context, another participant commented that the societal judgement and the legal age for accessing certain SRHR services may not be congruent with cultural norms within MRY communities, causing conflict and confusion. She stated, *“The legal age is 16, but can a 16-year-old walk into a Priceline pharmacy and buy plan B without people [like] judging or questioning or [like] giving a double take…?" (Jas, F, Indian)*.

#### Religious beliefs

Religious beliefs are often intertwined with cultural beliefs and can also influence attitudes towards SRHR. For example, a participant reflected, *“I also think it transcends into conversations about reproductive health… And what are the options?? Is contraception an option? Like, am I having kind of healthy amounts of pain, or is it debilitating? Can I go to a doctor? Like, I think the whole kind of sexual and reproductive health is a taboo in, well, like, I’m South Indian, so in my culture and I was raised Christian, so that also, religiously, abortion is never on the table" (Angelene, F, South Indian).* Another added, *“But going to a Catholic school, when they touched on sex, it was all about just abstinence” (Taylor, F, Filippino).* Subsequently, *“It was very hush-hush growing up. Abstinence is the best solution to this, and that’s basically what we were taught in primary school” (Lupita, F, Fijian).* Reflecting on her religion, another MRY remarked*, “If someone was raped or something like that, … she’d still be seen as dirty or impure. Yeah, because… her virginity has been taken before marriage” (Darlene, F, Ghanian).* For MRY from a religious background, these teachings discourage premarital sex, contraception, and abortion, without proffering support for SRH incidences thus affecting their experiences and understanding of SRHR.

#### Moral boundaries

The data also highlight that moral boundaries within families and communities can affect how MRY perceive and navigate SRHR. A participant rebuts her family’s presumption that the use of contraception would encourage sexual practices: *“They think being on the pill will encourage me to have sex instead of making my own decisions, so they would take any alternative route besides the pill…” (Lepa, F, Fijian).* Another participant reflects on the advice she received, *“The safest way for you to not get a sexual disease is to just don’t have it (sex), rather than use a condom or take the pill, etcetera” (Monique, F, Bosnian).* This presumptive approach tends to rob MRY of agency and decision-making capacity, instilling fear and promoting punitive consequences.

#### Media and culture

The portrayal of sex, sexuality, and reproductive health in media and popular culture shapes attitudes and beliefs about SRHR. According to the findings, exposure to pornography can lead to skewed perceptions of sexual relationships, particularly if it is a primary source of sexual education, as highlighted by MRY: *“Yeah, they’re just aggressive. They’re just like hitting them. And they think that that’s what women like” (Monique, F, Bosnian).* If media and popular culture perpetuate harmful stereotypes or misinformation about SRHR, this can influence MRY’s understanding of and engagement with their SRHR.

## Discussion

Throughout the focus group discussions, participants were encouraged to articulate their understanding of sexual and reproductive health and explore their perceptions of rights pertaining to SRH. Additionally, they were asked to identify and discuss barriers they had personally encountered in relation to SRH and to propose potential strategies to address these gaps. Significantly, most participants were unfamiliar with or had not previously considered their rights associated with SRH. Many expressed difficulties in distinguishing between sexual health and reproductive health, leading to many often-inconclusive understandings. Despite this, there was considerable alignment in participants’ responses across different focus groups about the individual barriers they faced in relation to SRHR.

Furthermore, participants mostly agreed that discussions about sex are frequently stigmatised to the extent that they overshadow associated components, such as health and well-being. This consensus underscores the pervasive impact of stigma across various spheres of influence, including familial relationships, cultural norms, education, and religious institutions. In separate studies, Asnong et al., (2018) and Logie et al. (2019) corroborated the findings suggesting that negative community attitudes towards sexual activity and access to reproductive services can deter youth from accessing information about sexual health services, including HIV testing. They also found that traditional views and stigma surrounding SRH issues contribute to a knowledge gap on contraception and life skills necessary for making informed choices among MRY (Asnong et al., 2018; Logie et al., 2019).

Using the Bronfenbrenner socioecological framework, this study identified barriers that influence intergenerational exchanges as they relate to MRY. The following is an interpretation of the results in line with Brofrenbrenner’s socioecological framework, in relation to existing evidence. The following sections include recommendations for practice, research, policy, and theory.

### Microsystem Level

At the microsystem level of Bronfenbrenner’s socioecological theory(Bronfenbrenner, 1979), which focuses on individuals and their immediate surroundings, this study uncovered numerous barriers that significantly impact the SRHR of MRY. A central issue identified is the lack of awareness and education regarding SRHR. Many MRY come from different cultural backgrounds where open discussions about SRHR are discouraged or non-existent (Khan et al., 2022). This lack of dialogue can lead to misunderstandings and misconceptions about SRHR. For instance, it was noted that participants had an unclear understanding of critical concepts such as consent. Ussher et al. (2017), Pound et al. (2016) and Khan et al. (2022) attributed the misunderstanding of such fundamental concepts to a lack of sex education, which is concerning because it exposes MRY to potential SRHR violations (Khan et al., 2022; Pound et al., 2016; Ussher et al., 2017). In their qualitative synthesis of young people’s views on school-based sex and relationship education (SRE), Pound et al. (2016) found that schools often approach SRE in the same way as other subjects without acknowledging the sensitive nature of the topic. This lack of acknowledgment and appropriate education can contribute to MRY misconceptions about SRHR. Our findings therefore align with existing research that emphasises the role of comprehensive education in enhancing SRHR outcomes (Khan et al., 2022; Ussher et al., 2017).

The fear associated with the likely consequences of sexual activity was another crucial barrier that was identified. Such fear is often rooted in inadequate or fear-based sexual education and tends to discourage MRY from engaging in safe sexual practices. This finding is consistent with Haas et al. (2017) and Mittal et al. (2013) studies that fear-based education can be counterproductive in promoting safe sexual practices (Haas et al., 2017; Mittal et al., 2013).

Similarly, culturally or socially instigated feelings of shame, guilt, and blame regarding SRHR were also identified as significant deterrents in MRY accessing necessary services or openly discussing SRHR issues. Baigry et al.’s (2023) recent findings identify social stigma, fear, and shame as barriers to accessing SRH services for young people in various countries, including Kenya, Nigeria, Malaysia, Nepal, and Iran – a similar demography represented in our study. Socio-cultural norms were also found to hinder youth’s access to contraceptives and STI treatments (Baigry et al., 2023). Similarly, the study by James et al. (2020) highlights the association between stigma and healthcare utilisation, emphasising how stigma can lead to experiences of shame, guilt, and blame, ultimately deterring MRY from seeking adequate healthcare services (James et al., 2020). Thus, our findings resonate with global research pointing to the pervasive influence of cultural and societal norms in shaping MRY’s SRHR outcomes (Wado et al., 2020).

While MRY in this study expressed concerns about their lack of knowledge about SRHR resulting in the inaccessibility of relevant services, Josefsson et al.’s (2019) study explores a separate challenge. Their study in Sweden revealed that students and professionals in various fields, including healthcare and social work, reported inadequate training and a lack of competence in SRHR (Josefsson et al., 2019). The lack of knowledge amongst professionals can intersect with the lack of knowledge amongst MRY can lead to dire consequences for MRY’s SRH. MRY unfamiliarity with the healthcare system and the geographically disadvantaged position of SRH centres, especially in Greater Western Sydney, add to the challenges. Such challenges highlight the crucial role of easily accessible and well-funded health services in promoting SRHR.

In addition, the influence of AOD on decision-making concerning SRHR emerged as a critical concern. Among other research, Horyniak et al.’s (2016) work has shown that forced migrants, including refugees, may be at risk for substance use as a coping mechanism for traumatic experiences, mental health disorders, acculturation challenges, and social and economic inequality (Horyniak et al., 2016). However, substance use can impair clear thinking and lead to unsafe sexual practices such as inconsistent condom use and multiple high-risk partners among MRY, echoing broader research linking substance use to poor SRHR outcomes (Elkington et al., 2009; Horyniak et al., 2016; Li et al., 2013; Mengesha et al., 2017).

Thus, microsystem-level barriers emphasise the complexity and multifaceted nature of the SRHR issues faced by MRY (Corosky & Blystad, 2016; Tirado et al., 2022). They underline the need for interventions to be person-centred, considering individual experiences, feelings, and immediate contexts. It corroborates Bronfenbrenner’s assertion of the microsystem’s significant role in shaping an individual’s experiences. This insight is essential for designing effective SRHR interventions for MRY, underscoring the need for comprehensive education, fear reduction, de-stigmatisation, adequate service accessibility, and AOD harm reduction strategies (Mengesha et al., 2017; Zelalem B Mengesha et al., 2018; Tirado et al., 2020).

#### Recommendations for Practice

MRY’s SRHR is subject to many socioecological influences within the context of Bronfenbrenner’s socioecological framework across system levels (Bronfenbrenner, 1979). To address these multilevel challenges, an evidence-based approach incorporating recommendations from the MRY is essential.

At the microsystem level, interpersonal relationships and immediate surroundings considerably affect MRY’s understanding of their SRHR. The absence of support from friends, parents, and teachers due to stigmatisation of these topics often leads to misinformation or a lack of information (Mengesha et al., 2017). This underscores the necessity of professional support and the creation of strong support networks, such as peer groups, school-based groups, and culturally appropriate networks, where comprehensive sex education programs are available (Mengesha et al., 2017). To tackle this issue, the SRHR of MRY must be established and promoted through a blend of system improvements and targeted services (Ussher et al., 2017). Service providers should create culturally safe health promotion strategies that are sensitive to the unique challenges faced by MRY, and integrate sexual health promotion into the early resettlement process (Fair et al., 2021; Tirado et al., 2022). Such strategies should encompass clear information regarding consent and other SRHR concepts. By offering comprehensive sex education and encouraging open discussions about consent, service providers can empower MRY to navigate their SRHR safely and with understanding (Fair et al., 2021).

To implement these strategies, professionals who interact with MRY should be trained in cultural safety and competency to adequately understand and support MRY’s unique SRHR needs (Botfield et al., 2016; Zelalem Birhanu Mengesha et al., 2018). Furthermore, comprehensive and culturally safe sex education programs should be developed to address the misinformation or lack of information MRY face (Roberts et al., 2017).

#### Mesosystem level

Mesosystem-level factors illustrate the complex web of socio-environmental barriers influencing MRY’s SRHR, highlighting the importance of comprehensive, culturally safe, and MRY- oriented interventions (Bronfenbrenner, 1979). At the mesosystem level, the interplay between various microsystems such as family, school, peer groups, and healthcare professionals plays a significant role in shaping the SRHR of MRY (Bronfenbrenner & Morris, 2007). These findings suggest the presence of numerous socio-ecological factors that affect MRY SRH outcomes.

One example is family conflict due to generational beliefs and cultural differences in SRH perspectives, which can present a significant hurdle. This conflict can hinder open discussions, which are crucial for imparting accurate and comprehensive SRHR information. This lack of open dialogue within families may lead to potentially harmful misconceptions about SRHR, with repercussions for MRY’s health and wellbeing (Huang et al., 2022). Huang et al.’s (2022) work suggested that a negative experience due to sexual stigma promotes secrecy, hindering help-seeking among minority groups; this aligns with our findings. Equally, Schaaf and Khosla (2021) examined the effectiveness of a culturally sensitive parent-adolescent communication intervention in promoting sexual health communication and reducing sexual risk behaviours in youth from low and middle income countries (Schaaf & Khosla, 2021). Their study found that the intervention significantly increased parent-adolescent communication about SRH topics and improved adolescents’ agency and knowledge of SRH. Miller et al. (2019) research had similar sexual health outcomes, including increased condom use and decreased risky sexual behaviours among African-American adolescents. Effectively, culturally sensitive and psychologically safe parent-adolescent communication initiatives are needed to address this barrier and promote accurate and comprehensive SRHR information and agency among MRY (Huang et al., 2022; Miller et al., 2019).

Another critical mesosystem-level barrier is the stigma surrounding sexual experiences, which can exacerbate social isolation among MRY. This finding aligns with Adinew et al. (2013) and Logie et al.’s (2019) work, which found that negative community attitudes towards sexual activity and access to reproductive services can deter youth from accessing information about SRH services, including HIV testing (Adinew et al., 2013; Logie et al., 2019). Additionally, Napier-Raman et al.’s (2023) systematic review conducted in Australia found that MRY face similar barriers to accessing services and care for their SRH. This indicates that stigma and other factors can contribute to social isolation and limited access to support for SRHR and peer education (Logie et al., 2019; Napier- Raman et al., 2023). Peer interactions are essential for sharing and learning experiences, particularly regarding sexual health. Our finding highlights the importance of creating safe spaces for these discussions, emphasising the importance of age, gender and culturally tailored programs to engage MRY in SRHR conversations (combating social stigmas), research and programming (Logie et al., 2019).

The gender knowledge gap, or the disparity in understanding and knowledge about sexual health between genders, has emerged in our study as a potential contributor to unsafe sexual practices and reinforced gender inequality (Endler et al., 2021; Gonzales et al., 2016; Torke & Carnahan, 2017; Veenstra, 2011). This gap is exacerbated by the portrayal of unrealistic and harmful views of sexual health and relationships in media and pornography (Miller et al., 2019). Miller et al.’s (2019) study shows that frequent pornography use is associated with sexual dissatisfaction and a greater preference for porn-like sex (Sommet & Berent, 2022; Wright et al., 2021). Our findings align with Miller et al. (2019), among other studies, that the use of pornography is linked to poor sex practices, violence and relationship dissatisfaction among MRY (Sommet & Berent, 2022; Wright et al., 2021).

In tandem, experiences or exposure to sexual violence, a grave concern at the mesosystem level, can profoundly impact SRHR by inducing trauma and impacting future sexual behaviour (Kalra & Bhugra, 2013). Kalra and Bhugra (2013) argue that migrants and refugees, particularly women, adolescents, and children, often experience physical and/or sexual violence along the migratory route, and these experiences continue to have a psychological impact on migrants and refugees’ lives in their destination country and are key factors preventing access to appropriate health and social care (Kalra & Bhugra, 2013; Tirado et al., 2020). Our findings reveal that the significant people in MRY’s lives may further amplify these traumatic experiences and deepen the social and psychological impact. This is supported by Keygnaert et al.’s (2015) research (on sexual violence among refugees, people seeking asylum, and undocumented migrants in various countries) which demonstrated the frequent co-occurrence of sexual violence with physical, psychological, and socio-economic forms of violence (Keygnaert & Guieu, 2015). Consequently, MRY victims disproportionately experience mental health challenges, including depression shaped by trauma, poverty, and elevated exposure to sexual and gender-based violence (Logie et al., 2019). Substance use and depression often co-occur among forced migrants, further exacerbating their mental health disparities (Logie et al., 2019).

Based on our findings, the lack of confidentiality and trust at the mesosystem level can also foster feelings of isolation and anxiety among MRY. This claim is supported by several studies (Lynch et al., 2021; Maheen et al., 2021; McCann et al., 2016; Zelalem Birhanu Mengesha et al., 2018; Tirado et al., 2022). The absence of trusted guidance or representation can complicate MRY’s navigation of SRHR, particularly considering cultural safety and fear of judgement. Tirado et al. (2022) explore a parallel concern for MRY where an interpreter is required, highlighting the interpreters’ judgment and confidentiality when discussing sensitive SRHR subjects with the MRY. Reliance on untrained interpreters for discussing SRHR may result in language discordance and difficulties for clients to gain comprehensive and accurate health-related information. The need for confidential services in SRHR is underscored, particularly those specifically designed for MRY (Lynch et al., 2021).

Furthermore, insensitivity towards SRHR from educators, service providers, or familial adults can exacerbate the stigma and fear associated with SRH, thereby depleting MRY agency and help-seeking capacity (Cohodes et al., 2021; Tirado et al., 2022). Several studies have highlighted the positive impact of safe and sensitive SRHR education and services on young people’s well-being and SRH attitudes. For example, a study conducted by Tirado et al. (2022) in Sweden found that there are fragments in SRH services for young migrants, including a lack of knowledge about SRHR among migrant youth, language and communication barriers, and a lack of structure needed to build dependable services that go beyond one-time interventions. Similarly, a study by Cohodes et al. (2021) emphasised the importance of providing youth-friendly SRH services that are respectful, confidential, and non-discriminatory. In their Inuit youth research conducted in Canada, Corosky and Blystad (2016) found that youth face significant barriers to SRHR care and support, including a lack of trust in support workers, stigma and taboos surrounding SRHR topics, and feelings of powerlessness. These barriers particularly affect female youth, making it even more crucial to create an environment where MRY feel comfortable seeking help (Corosky & Blystad, 2016; Davis et al., 2017). The need for sensitivity in these areas is paramount to ensure that MRY feel comfortable and empowered to seek adequate SRH support.

#### Recommendation for Practice

At the mesosystem level, a lack of interconnectedness between microsystem entities (such as family, school, and community) leads to gaps in knowledge, understanding, and access to services for SRHR (Botfield et al., 2017; Corosky & Blystad, 2016; Davis et al., 2017). Youth services can liaise with other community managed organisations and the department of education (through the school system) to facilitate MRY groups and workshops. Introducing MRY groups and workshops can serve as bridging points between these entities. This can foster a more comprehensive understanding, and enable MRY to navigate SRHR services more effectively. These MRY groups can serve as platforms where the importance of SRHR awareness, knowledge, and value clarification are discussed at the individual and community levels to promote a supportive attitude towards SRHR.

#### Recommendation for Research

Research has indicated that early formative environments, such as schools and communities, profoundly influence the attitudes and behaviours of youth regarding SRHR (Mmari et al., 2014; Mulubwa et al., 2020; Zulu et al., 2018). Collaboration between these entities could foster a nurturing environment, instilling respectful and inclusive SRHR attitudes from an early age. Thus, future research should aim to develop and evaluate collaborative programs involving various stakeholders in SRHR education and to understand their impact on the well-being of MRY. In addition, future research should investigate the effectiveness of peer-led education strategies in promoting healthy SRHR behaviours and determine the factors contributing to their success. Based on the insights from this study, peer groups are often a significant source of information and influence among youth. Therefore, understanding peers’ roles in shaping SRHR behaviours and beliefs can inform the development of peer-led interventions.

#### Exosystem Level

The barriers presented at the exosystem level, such as lack of appropriate professional support, language barriers, policy impacts, absence of comprehensive SRHR education in the curriculum, and lack of access to services, reflect the complexity and interconnectedness of factors emphasised by Bronfenbrenner’s socioecological model (Cohodes et al., 2021; Corosky & Blystad, 2016; Endler et al., 2021; Mengesha et al., 2017; Tirado et al., 2020; Tirado et al., 2022). These barriers hinder the SRHR of MRY and are influenced by various structural and systemic factors (Tirado et al., 2020).

The absence of professionals (such as culturally safe SRHR counsellors, GPs, teachers and social workers) to guide MRY through SRHR issues, has significant implications. The research conducted by Tirado et al. (2022) regarding the barriers faced by migrant youth in accessing SRH services in Sweden supports our findings. The study highlighted the importance of improving healthcare providers’ awareness and culturally safe SRHR services for migrant youth. In line with our findings, Tirado et al. (2022) and Aibangbee et al. (2023) note that navigating SRHR within new societies can be particularly challenging for MRY without professional support that understands the unique needs and complex issues MRY navigate daily.

Language barriers, a well-documented issue among migrant and refugee populations, further intensify the challenge, inhibiting MRY’s ability to understand and engage with SRHR services (Aibangbee et al., 2023; Botfield et al., 2017; Gray et al., 2021; Mengesha et al., 2017). This communication difficulty complicates MRY’s chosen acculturation strategy and most likely results in social isolation and misunderstanding of crucial information about SRHR, exacerbating health risks (Gray et al., 2021).

The impact of policies on MRY’s SRHR is another factor at the exosystem level (Tirado et al., 2020). Policies that do not acknowledge cultural implications or are inclusive of the unique needs of MRY can create additional barriers to their access to SRHR (Tirado et al., 2020). For example, taxing menstrual health products can further emphasise socio-economic inequities and impact MRY’s SRHR (Tirado et al., 2020). Similarly, migration-related trauma complicates MRY’s response to the system. For example, Cohodes et al. (2021) study examined the effects of migration-related trauma on the mental health of young migrants emigrating from Mexico and Central America to the United States. This buttresses our findings on the impact of structural factors on the well-being of MRY and their responsiveness to SRHR.

The lack of comprehensive SRHR education in the school curriculum reiterated at the microsystem level, is a systemic educational barrier that negatively impacts MRY’s understanding of and access to SRHR information and services (Aibangbee et al., 2023; Napier-Raman et al., 2023; Villa-Torres & Svanemyr, 2015). Various research including Villa-Torres and Svanemyr’s (2015) work consistently demonstrates the significant impact of SRH education on promoting safer sexual practices and mitigating health risks, as evidenced by our findings. The outcome underscores the importance of educational reform to include a comprehensive SRHR education, considering the unique needs and practical implications for MRY.

Furthermore, limited access to services, compounded by issues such as stigma, lack of knowledge about service availability, and logistical challenges, suggests significant environmental barriers at the exosystem level (Gray et al., 2021). These barriers align with Bronfenbrenner’s model, which identifies the systems and structures within the broader environment as significant influences on individual development and behaviour (Bronfenbrenner, 1979).

These findings highlight the critical need to address the various barriers at the exosystem level that affect MRY’s SRHR. They underline the importance of integrated and multilevel approaches that address systemic issues in education, policy, and service provision, in line with Bronfenbrenner’s socioecological theory (Corosky & Blystad, 2016). Furthermore, they point to the necessity of cultural safety and inclusive practices that consider the unique needs and experiences of MRY (Tirado et al., 2020).

#### Recommendations for Practice

Our study observes that existing SRH services may not fully resonate with the distinct needs and experiences encountered by MRY in various regions, including Greater Western Sydney. This gap signifies the pressing need to restructure and realign these services to be more inclusive and responsive (Azami-Aghdash et al., 2015).

Several strategies, including language-accessible resources, can be initiated to bridge this gap. It is critical to offer SRHR materials and resources in multiple languages. This approach ensures that relevant communication and language support are improved to mitigate language barriers that hinder access to SRHR services. Additionally, introducing multilingual staff in health services can provide more personalised assistance, fostering a sense of safety and understanding for MRY during their health visits (Tirado et al., 2022). Also, developing MRY-friendly platforms (including in prevailing social media platforms) where they can provide direct feedback and present ideas in a safe and collaborative space, can foster an inclusive healthcare environment.

Another crucial step in addressing MRY’s needs is implementing community advertising initiatives. This is operationalised through collaboration with community leaders (including influencers and gatekeepers) and organisations to develop campaigns that resonate with MRY. The campaign can be designed to increase awareness about available SRHR services, encourage community dialogue on these topics and foster a supportive environment for MRY(Thornicroft et al., 2008; Zulu et al., 2018).

These recommendations are not without challenges. In agreement with Tirado et al. (2022) and VanderWielen et al.’s (2014) work, our findings reiterate concerns about the privacy, accuracy, and confidentiality for interpreters. These concerns further emphasise the need to train interpreters and multilingual workers to have a broader understanding of SRHR topics to ensure the effectiveness and long-term success of these strategies (Tirado et al., 2022; VanderWielen et al., 2014).

#### Recommendation for Research

The insights from this study identified the importance of intergenerational dialogues in destigmatising and normalising SRHR conversations. To examine this further, future research could focus on designing and evaluating interventions to test and promote SRHR dialogues within MRY communities. Given the small representation of male participants in the study, future research could also consider MRY gender-specific SRHR exploration. This aligns with the findings of Ruane- Mcateer et al. (2019) who emphasised the importance of gender-transformative programming to engage men and boys in SRHR decision-making and improve SRHR outcomes (Ruane-McAteer et al., 2019).

Furthermore, addressing the unique needs of MRY with disabilities is crucial for ensuring inclusive SRHR policies and programs. In line with the study by Hameed et al. (2020), future studies should focus on developing evidence-based policies and interventions to support MRY with disabilities, including identifying their specific SRHR barriers and effective strategies for overcoming them(Hameed et al., 2020). Also, the study by Scherer et al. (2021) emphasised the importance of coordination, efficiency, and accountability in disability-inclusive programs, which can also be applied to SRHR interventions for MRY with disabilities (Scherer et al., 2021).

#### Recommendation for Policy

Policies can be formulated to develop culturally safe and inclusive healthcare systems that address the unique needs and barriers MRY face. Policies should include implementing cultural safety training as a mandatory component of continuous professional development for healthcare providers. In their work, Curtis et al. (2019) highlight that cultural safety training helps healthcare providers understand and address the cultural, social, and historical factors that influence the health outcomes of MRY. It promotes self-reflection, awareness of power dynamics, and the provision of care that is respectful, safe, and responsive to the needs of diverse populations (Curtis et al., 2019; Karatay et al., 2016; Lonne et al., 2020).

Policies should prioritise the allocation of resources for future research and development to better understand the specific SRHR needs and prevailing challenges faced by MRY. This research should adopt a bottom-up and collaborative demography-led approach, involving the active participation of MRY and their communities similar to the methodology applied in our research (Dune et al., 2018; Gifford et al., 2019). Evidence-based policies and programs can then be formulated based on the findings of this research, ensuring that they are tailored to the unique needs and experiences of MRY (Brooks-Cleator et al., 2018; Gifford et al., 2023).

Policies should promote inter-agency collaboration between local health districts and community-managed organisations to ensure that services and programs addressing the SRHR of MRY are coordinated, comprehensive, and effective. Roseby et al. (2019) opine that collaboration which begins at the local level and involves the active participation of MRY and their communities is mostly effective. Therefore, through collaborative efforts, different stakeholders can pool their resources, expertise, and knowledge to develop and implement holistic approaches to SRHR that address the social determinants of health and promote health equity (Gifford et al., 2019; Roseby et al., 2019).

By implementing these recommendations, policymakers can contribute to the development of culturally safe and inclusive healthcare systems that address the unique needs and barriers faced by MRY in accessing SRHR services. These policies can help reduce health disparities, promote health equity, and ensure MRY’s agency.

#### Macrosystem level

At the macrosystem level, several cultural and societal factors play a role in shaping MRY’s SRHR. These factors reflect Bronfenbrenner’s socioecological theory’s outermost layer, which includes the overarching patterns of a given culture or subculture (Bronfenbrenner, 1979). The influence of these larger social systems on MRY’s SRHR can manifest in nuanced and complex ways. This research findings reveal the strong influence of cultural and societal norms on MRY’s SRHR. These norms can stigmatise discussions about SRHR, leading to limited access to vital information and resources (Asekun-Olarinmoye et al., 2014; Dune et al., 2017; Rawson & Liamputtong, 2010). This cultural influence aligns with Bronfenbrenner’s emphasis on the macrosystem’s role in shaping behaviours and experiences, indicating a need for interventions that challenge harmful norms and promote SRHR-friendly cultural shifts (Bronfenbrenner, 1979; Mulubwa et al., 2020).

Religious beliefs, which are often intertwined with cultural norms, can significantly influence SRHR attitudes. Mpofu’s (2018) research on health and wellbeing among religious adherents in Zimbabwe found that religious beliefs potentially expose women and children to health risks. They potentially pose barriers to services such as contraception and abortion and limit discussions about SRH (Amroussia, 2022; Mpofu, 2018; Mpofu et al., 2014). Our study underscores the need for culturally safe and religiously sensitive health education and services that respect religious beliefs and ensure comprehensive access to SRHRs (Mpofu et al., 2014).

Our study also highlights how moral boundaries within families and communities complicate MRY’s SRHR. These moral boundaries can inadvertently perpetuate fear and limit MRY’s agency in decision-making about their SRHR. This observation aligns with Bronfenbrenner’s macrosystem model, which identifies societal norms as significantly influencing individual behaviours (Bronfenbrenner, 1979; Mbarushimana et al., 2022).

Moreso, our findings reveal that portraying sex, sexuality, and reproductive health in media and popular culture plays a significant role in shaping young people’s attitudes towards SRHR. Aylward & Halford’s (2020) research, which reiterates the United Nations’s stance on the rights of children to access SRH services and evidence-based education on human sexuality, tends to agree with Kwankye and Augustt (2013) research hypothesis. They hypothesised that exposure to the media influences young people to adopt positive SRH behaviour. While partly true, Kwankye and Augustt’s (2013) overall findings do not consistently show statistically significant associations between media exposure and reproductive health behaviour (Kwankye & Augustt, 2013). In alignment with our findings, it indicates that negative or unrealistic representations can lead to misunderstandings of sexual relationships and contribute to inaccurate and misinformative portrayals in media (Aylward & Halford, 2020). According to our findings, these portrayals can also inadvertently result in a trado- cultural insistence on shielding young people from over-sexualised Western society.

Conversely, legal and political structures significantly impact MRY’s access to SRHR services, as supported by Gazard et al. (2018). For instance, restrictive immigration policies can limit healthcare access for MRY, emphasising how migration status intersects with SRHR access. Godwin et al. (2017) note that a range of other factors such as ethnicity and socioeconomic status further complicate this intersectionality. Discriminatory experiences arising from these intersecting identities can increase the need for SRHR services and create disparities in their utilisation (Goodwin et al., 2017). Our findings stress the importance of considering these multi-layered discrimination factors and advocating for an intersectional approach in research on health service use. The conflict between legal age and cultural norms is another example, further illustrating the intricate interplay between macrosystem factors and MRY’s experiences with SRHR services (Gazard et al., 2018).

Thus, the findings revealed that the barriers identified at the macrosystem level emphasise the critical need for comprehensive, culturally safe, and accessible SRHR services and education. Furthermore, they highlight the need for larger societal changes, including legal and policy reforms, and shifts in cultural and societal norms in line with Bronfenbrenner’s socioecological theory.

#### Recommendations for Practice

At the macrosystem level, societal norms, values, and laws facilitating MRY’s SRHR cannot be realised without effective intergenerational and intercultural dialogue (Beauregard et al., 2019). Our findings revealed that MRY often navigate conflicting cultural norms and stigmas, which can exacerbate their SRHR challenges. Therefore, intergenerational dialogues are crucial in destigmatising and normalising discussions about SRHR. These dialogues facilitate the exchange of knowledge and experiences between different generations, leading to a better understanding and acceptance of SRHR issues (Branquinho et al., 2022; Ogbe et al., 2018; Schmitt et al., 2015). Drawing from Schmitt et al.’s (2015) analysis of SRHR in post-Soviet society, intergenerational dialogue contributed to more effective use of older people’s potential for generativity. Such strategy provides an opportunity for joint agenda-setting and policy framing, and advocating prioritising purposive SRHR issues (Ogbe et al., 2018).

Implementing intergenerational dialogues to address MRY’s SRHR challenges at the macrosystem level would necessitate a multifaceted approach. An initial step could include organising community-based forums or workshops (‘town hall meetings’) where different generations can come together to discuss SRHR issues. These gatherings would provide a platform for the older migrant and refugee generation to share historical perspectives on SRHR and for MRY to voice their current concerns and experiences. Incorporating MRY’s recommendations into practice can help ensure that they have the knowledge, resources, and support they need to make informed decisions about their SRHR.

Secondly, schools and universities could incorporate SRHR dialogues into their curricula or extracurricular programs, perhaps as part of a larger public health education initiative (Botfield et al., 2017). This could include guest lectures from older community members or SRHR health providers.

Also, media platforms can be used to normalise and destigmatise conversations around SRHR. This could include interviews, podcasts, or social media campaigns featuring intergenerational dialogues on SRHR topics. Furthermore, engagement with religious leaders and institutions could be particularly influential in shifting cultural norms. This may involve educating religious leaders on the importance of SRHR and equipping them to facilitate dialogues within their communities. Conversely, Mpofu et al. (2017) stress that harmonising the religious and cultural aspects of SRHR policies is essential. This can be facilitated through interfaith dialogues to cultivate understanding and collaboration with cultural leaders, thereby promoting a more receptive environment.

Equally, older individuals can be trained to serve as SRHR advocates, drawing upon their life experiences to enrich the dialogue. Similarly, youth could be trained in effective communication and advocacy skills to better express their SRHR needs and perspectives.

#### Recommendations for Theory

This research project has used Bronfenbrenner’s socioecological theoretical framework (1979; 2007) as a lens to investigate and understand the barriers MRY encounter in relation to their SRHR. While this study reveals how each socioecological system is interlinked, it does not incorporate a significant system— the chronosystem— initially omitted from Bronfenbrenner’s model (1979), but later included to account for environmental changes over time (Dulin et al., 2018).

The chronosystem considers significant life transitions, socio-historical events, and environmental changes over time, which can significantly affect an individual’s development. The inclusion of this perspective is particularly relevant when examining MRY experiences. Migration itself is a significant life event, and the time since migration, the age at migration, and the historical and sociopolitical context of the migration period can all significantly impact MRY’s SRHR. Therefore, understanding how MRY’s SRHR evolves over time in response to changing personal circumstances and broader sociocultural environments can provide important insights into the long-term impacts of current interventions and identify areas where additional support may be needed.

While the socioecological model offers invaluable insights into the realities of individuals and communities, it is essential to guard against assumptions and biases that could emerge inadvertently. For instance, not all MRY encounter poor SRHR conditions or lack safe microsystems for open discussions on SRHR. Thus, the socioecological model should be viewed as a tool for understanding the complexities within individual experiences and communities, rather than a mechanism for making extensive generalisations.

Moreover, a more comprehensive theoretical approach requires the inclusion of other vulnerable MRY demographics. This can span from the LGBTQI+ community to MRY living with disabilities. Their unique experiences could illuminate different dimensions of the challenges faced and opportunities that can be harnessed, thereby deepening and enriching the existing theoretical framework (Herrick et al., 2013; Prather et al., 2016).

Adopting an intersectional lens within the socioecological model can further illuminate how various social identities such as race, gender, and sexual orientation intersect at the micro, meso, exo, and macro levels. Such intersections shape the SRHR experiences of MRY in profound ways and acknowledging them can ensure the development of strategies that are more responsive to these intersectional experiences.

#### Strengths & Limitations

This research project was designed to examine the SRHR experiences and challenges of MRY in Australia’s Greater Western Sydney, and it succeeded in gathering rich and nuanced data. A key strength of this study was its utilisation of the socioecological model, which provided a robust theoretical framework for examining SRHR experiences from a multidimensional perspective. The framework allowed for a comprehensive understanding of MRYs’ experiences across different environmental systems and the interplay between these systems.

In addition, applying the PAR methodology provided a platform for participants to actively engage in the research process and contribute to understanding their individual experiences. This enhanced the authenticity and depth of the study, capturing the lived experiences of MRYs from their perspective. The diversity in the participants’ cultural backgrounds and experiences further enhanced the richness of the data and the insights drawn.

However, despite these strengths, the study has some limitations. The sample had an overrepresentation of female participants out of those who submitted their demographic information, comprising approximately 70%, which might limit the applicability of the findings to the broader population of MRYs in Australia. The COVID-19 pandemic significantly influenced the design and execution of the study, which led to most of the focus group sessions being conducted online. This change might have affected the depth of interaction, even though it provided a safe platform for MRY to participate in the study. Technical issues such as internet connectivity hindered full participation by a small number of participants but did not significantly impact the data. Furthermore, the pandemic situation meant that participants’ attendance was lower than initially envisaged at each focus group, resulting in an increase of completed focus groups by 80% to achieve data saturation. These changes potentially affected the generalisability of the findings to non-pandemic times.

Thus, future research should seek to address these limitations, possibly by aiming for a more balanced gender representation and accounting for the influence of exceptional circumstances, such as a global pandemic, on the study’s outcomes. Despite these limitations, the findings offer valuable insights into the SRHR experiences of MRYs and provide a solid foundation for further research in this field.

## Conclusion

In conclusion, this study illuminates the complex and multidimensional experiences of MRY’s SRHR in Australia. Employing Bronfenbrenner’s socioecological model affords an understanding of the nuanced influences of various environmental systems on MRY’s SRHR. These influences range from microsystem factors, such as family dynamics, to broader macrosystem influences, such as national health policies, with each level integral to shaping MRYs’ SRHR experiences.

Empathy and analytical rigour formed the basis for understanding the myriad barriers MRY face in relation to their SRHR. These barriers are often deeply rooted in socio-cultural nuances, economic disparities, and sometimes, policy inadequacies that create a complex web of challenges that MRY face in accessing adequate SRH services and resources. It becomes critical, therefore, to dissect the underpinnings of these barriers — to understand why they exist or do not exist — in a bid to formulate evidence-based strategies to address them. Recognising the depth and breadth of the findings in this study, we acknowledge that this work is preliminary considering the intricate dynamics at play. Recognising the depth of work, we anticipate delving further into this vital study area. A forthcoming paper will be dedicated to articulating the facilitators and solutions proposed directly by the MRY, harnessing their firsthand insights and experiences to pave the way for more grounded, inclusive, and effective interventions in this space. This progressive step aims not only to illuminate the intricacies of these barriers but also to actively engage in a dialogue that fosters solutions, resilience, and empowerment among the MRY community in Australia.

Valuable insights were obtained from the participants using the PAR methodology. This approach emphasised the importance of engaging directly with communities and accurately capturing lived experiences. The recommendations provided for practice, research, policy and theory offer a path forward for enhancing the SRHR experience of MRYs. However, limitations of the study, such as the gender imbalance in the sample and the potential impacts of the COVID-19 pandemic on research execution and outcomes, warrant acknowledgement. Future research must strive to address these limitations to ensure wider applicability of the findings.

Overall, this study signifies that addressing SRHR challenges confronting MRY is a multilevel and complex task. Collective action across multiple levels of society, from individual families to broader socio-political contexts, is required. The findings from this study provide a crucial stepping stone towards expanding knowledge, policymaking, and practice to support MRYs’ SRHR agency and decision-making in Australia.

## Data Availability

Availability of data and materials: The dataset generated and/or analysed during the current study is available under restricted access. The data is located in the Research Data Australia portal at https://doi.org/10.26183/2x5y-v748. For more information or potential collaboration, please contact the corresponding author.

## Acknowledgement

The authors declare that this study received funding from the Australian Research Council (grant number: DP200103716) and the Western Sydney University Central Funding covering the article processing charge. The funders were not involved in the study design, collection, analysis, interpretation of data, the writing of this article or the decision to submit the manuscript for publication.

## Availability of data and materials

The dataset generated and/or analysed during the current study is available under restricted access. The data is located in the Research Data Australia portal at https://doi.org/10.26183/2x5y-v748. For more information or potential collaboration, please contact the corresponding author.

## Declaration of interest statement

The authors declare that the research was conducted in the absence of any commercial or financial relationship that could be construed as a potential conflict of interest.

## APPENDIX A Participant Demographics

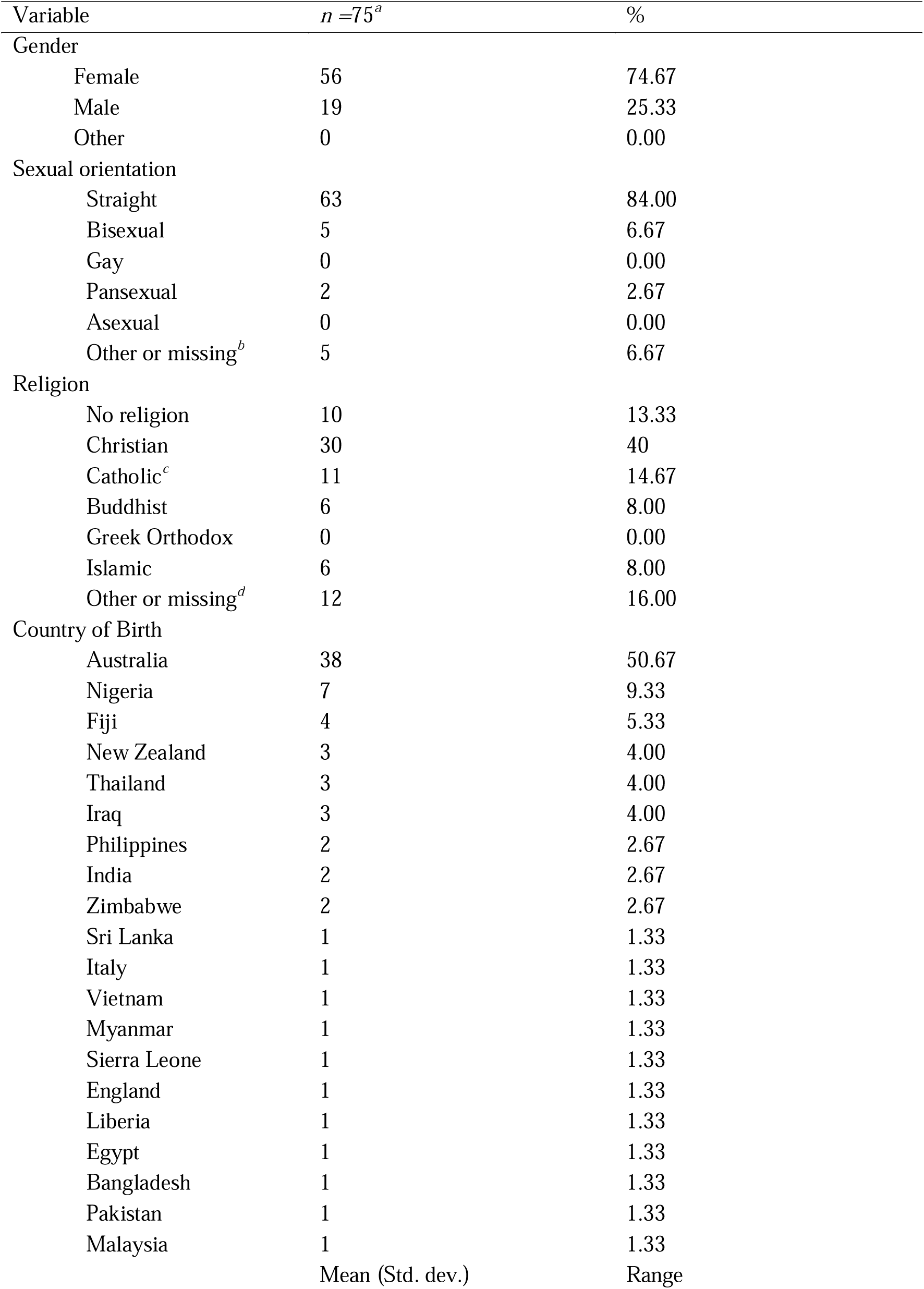
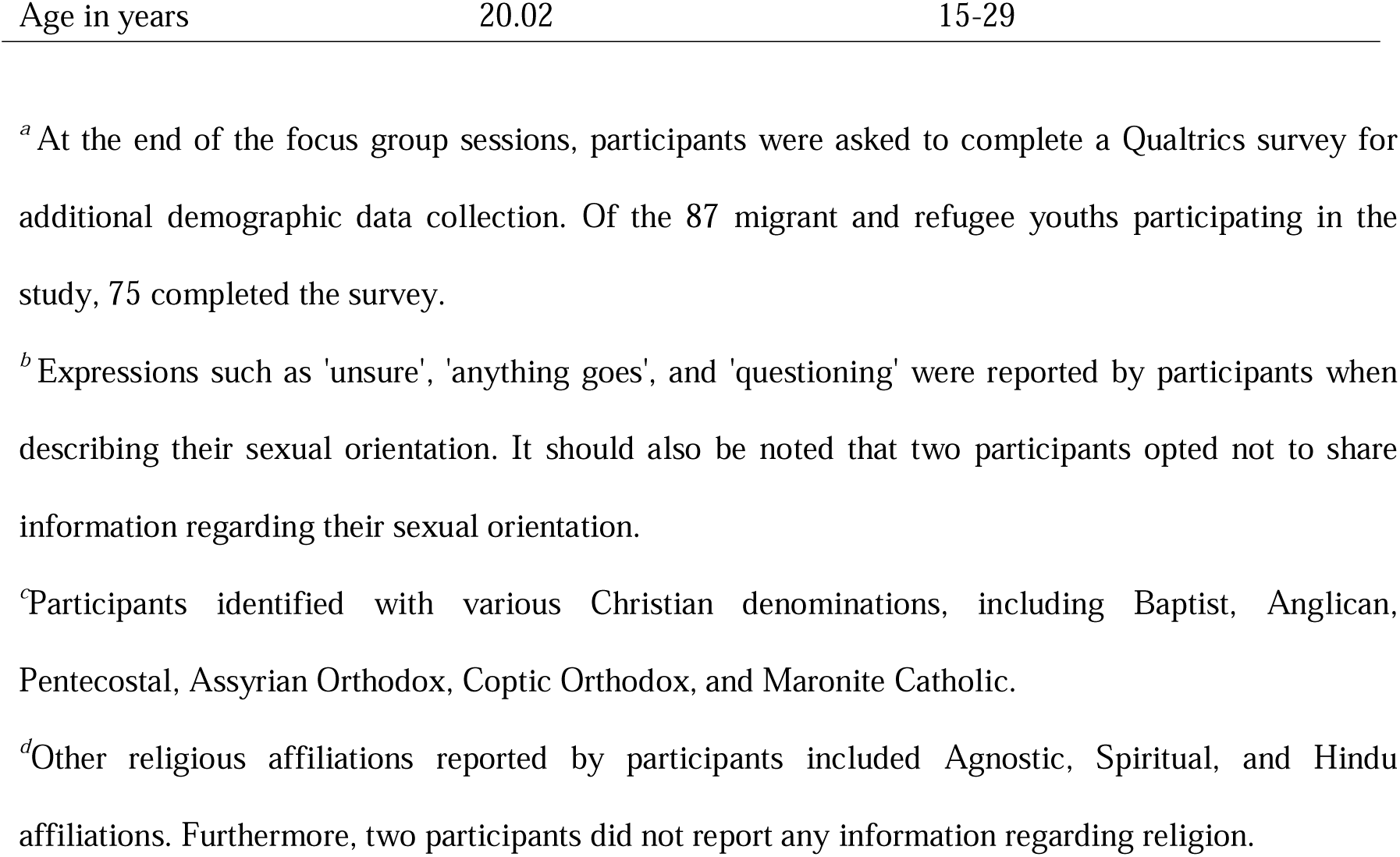

Participant Demographics
| Variable | *n* = 75<sup>a</sup> | % |
| --- | --- | --- |
| Gender |  |  |
| Female | 56 | 74.67 |
| Male | 19 | 25.33 |
| Other | 0 | 0.00 |
| Sexual orientation |  |  |
| Straight | 63 | 84.00 |
| Bisexual | 5 | 6.67 |
| Gay | 0 | 0.00 |
| Pansexual | 2 | 2.67 |
| Asexual | 0 | 0.00 |
| Other or missing<sup>b</sup> | 5 | 6.67 |
| Religion |  |  |
| No religion | 10 | 13.33 |
| Christian | 30 | 40 |
| Catholic<sup>c</sup> | 11 | 14.67 |
| Buddhist | 6 | 8.00 |
| Greek Orthodox | 0 | 0.00 |
| Islamic | 6 | 8.00 |
| Other or missing<sup>d</sup> | 12 | 16.00 |
| Country of Birth |  |  |
| Australia | 38 | 50.67 |
| Nigeria | 7 | 9.33 |
| Fiji | 4 | 5.33 |
| New Zealand | 3 | 4.00 |
| Thailand | 3 | 4.00 |
| Iraq | 3 | 4.00 |
| Philippines | 2 | 2.67 |
| India | 2 | 2.67 |
| Zimbabwe | 2 | 2.67 |
| Sri Lanka | 1 | 1.33 |
| Italy | 1 | 1.33 |
| Vietnam | 1 | 1.33 |
| Myanmar | 1 | 1.33 |
| Sierra Leone | 1 | 1.33 |
| England | 1 | 1.33 |
| Liberia | 1 | 1.33 |
| Egypt | 1 | 1.33 |
| Bangladesh | 1 | 1.33 |
| Pakistan | 1 | 1.33 |
| Malaysia | 1 | 1.33 |
|  | Mean (Std. dev.) | Range |
| Age in years | 20.02 | 15-29 |

<sup>a</sup> At the end of the focus group sessions, participants were asked to complete a Qualtrics survey for additional demographic data collection. Of the 87 migrant and refugee youths participating in the study, 75 completed the survey.
<sup>b</sup> Expressions such as 'unsure', 'anything goes', and 'questioning' were reported by participants when describing their sexual orientation. It should also be noted that two participants opted not to share information regarding their sexual orientation.
<sup>c</sup> Participants identified with various Christian denominations, including Baptist, Anglican, Pentecostal, Assyrian Orthodox, Coptic Orthodox, and Maronite Catholic.
<sup>d</sup> Other religious affiliations reported by participants included Agnostic, Spiritual, and Hindu affiliations. Furthermore, two participants did not report any information regarding religion.

## Appendix B Focus group questions

1. What does the term sexual health mean to you?
2. What does the term reproductive health mean to you?
3. What are your human rights in relation to your sexual and reproductive health?
4. What helps you to maintain and protect your sexual reproductive health?
5. What stops you from being able to maintain or protect your sexual reproductive health?
6. What needs to be done differently in Western Sydney to address these SRHR gaps?

## Notes

### Competing Interest Statement

The authors have declared no competing interest.

### Author Declarations

Ethics Committee of Western Sydney University Human Research gave ethical approval (H13798) for this work.

## Reference

Adinew, Y. M., Worku, A., & Mengesha, Z. B. (2013). Knowledge of Reproductive and Sexual Rights Among University Students in Ethiopia: Institution-Based Cross-Sectional. *BMC International Health and Human Rights*. 10.1186/1472-698x-13-12

Aibangbee, M., Micheal, S., Mapedzahama, V., Liamputtong, P., Pithavadian, R., Hossain, Z., Mpofu, E., & Dune, T. (2023). Migrant and Refugee Youth’s Sexual and Reproductive Health and Rights: A Scoping Review to Inform Policies and Programs [Review]. International Journal of Public Health, 68. 10.3389/ijph.2023.1605801

Amroussia, N. (2022). Providing Sexual and Reproductive Health Services to Migrants in Southern Sweden: A Qualitative Exploration of Healthcare Providers’ Experiences. BMC Health Services Research. 10.1186/s12913-022-08967-3

Asekun-Olarinmoye, O. S., Asekun-Olarinmoye, E. O., Adebimpe, W. O., & Omisore, A. G. (2014). Effect of mass media and Internet on sexual behavior of undergraduates in Osogbo metropolis, Southwestern Nigeria. Adolescent health, medicine and therapeutics, 15-23.

Asnong, C., Fellmeth, G., Plugge, E., Wai, N. S., Pimanpanarak, M., Paw, M. K., Charunwatthana, P., Nosten, F., & McGready, R. (2018). Adolescents’ perceptions and experiences of pregnancy in refugee and migrant communities on the Thailand-Myanmar border: a qualitative study. Reproductive health, 15(1), 83. 10.1186/s12978-018-0522-7

Atuyambe, L., Kibira, S. P. S., Bukenya, J., Muhumuza, C., Apolot, R. R., & Mulogo, E. (2015). Understanding Sexual and Reproductive Health Needs of Adolescents: Evidence From a Formative Evaluation in Wakiso District, Uganda. Reproductive health. 10.1186/s12978-015-0026-7

Aylward, E., & Halford, S. (2020). How Gains for SRHR in the UN Have Remained Possible in a Changing Political Climate. Sexual and Reproductive Health Matters. 10.1080/26410397.2020.1741496

Azami-Aghdash, S., Ghojazadeh, M., Sheyklo, S. G., Daemi, A., Kolahdouzan, K., Mohseni, M., & Moosavi, A. (2015). Breast Cancer Screening Barriers From the Womans Perspective: A Meta-Synthesis. Asian Pacific Journal of Cancer Prevention. 10.7314/apjcp.2015.16.8.3463

Baigry, M. I., Ray, R., Lindsay, D., Kelly[Hanku, A., & Redman-MacLaren, M. (2023). Barriers and Enablers to Young People Accessing Sexual and Reproductive Health Services in Pacific Island Countries and Territories: A Scoping Review. PLoS One. 10.1371/journal.pone.0280667

Beauregard, C., Tremblay, J., Pomerleau, J., Simard, M., Bourgeois-Guérin, É., Lyke, C., & Rousseau, C. (2019). Building Communities in Tense Times: Fostering Connectedness Between Cultures and Generations Through Community Arts. American Journal of Community Psychology. 10.1002/ajcp.12411

Berry, J. W. (2005). Acculturation: Living successfully in two cultures. International journal of intercultural relations, 29(6), 697–712.

Botfield, J., Newman, C., & Anthony, Z. W. I. (2017). P4.51 Engaging young people from migrant and refugee backgrounds with sexual and reproductive health promotion and care in sydney, australia. Sexually Transmitted Infections, 93. 10.1136/sextrans-2017-053264.548

Botfield, J. R., Newman, C. E., & Zwi, A. B. (2016). Young people from culturally diverse backgrounds and their use of services for sexual and reproductive health needs: a structured scoping review. Sexual health, 13(1), 1. 10.1071/sh15090

Branquinho, C., Xavier, L. L., Andrade, C., Ferreira, T., Noronha, C., Wainwright, T., & de Matos, M. G. (2022). We Are Concerned about the Future and We Are Here to Support the Change: Let’s Talk and Work Together! *Children*, 9(10), 1574. https://www.mdpi.com/2227-9067/9/10/1574

Braun, V., & Clarke, V. (2019). Reflecting on reflexive thematic analysis. *Qualitative Research in Sport*, Exercise and Health, 11(4), 589–597. 10.1080/2159676X.2019.1628806

Bronfenbrenner, U. (1979). The ecology of human development: Experiments by nature and design. Harvard university press.

Bronfenbrenner, U., & Morris, P. A. (2007). The bioecological model of human development. Handbook of child psychology, 1.

Brooks-Cleator, L. A., Phillipps, B., & Giles, A. R. (2018). Culturally Safe Health Initiatives for Indigenous Peoples in Canada: A Scoping Review. Canadian Journal of Nursing Research. 10.1177/0844562118770334

Cavaleri, R., Mapedzahama, V., Pithavadian, R., Firdaus, R., Ayika, D., & Arora, A. (2021). Culturally and linguistically diverse Australians. In Diversity and Health in Australia (1st ed., pp. 145-159). Routledge. 10.4324/9781003138556

Chattu, V. K., & Yaya, S. (2020). Emerging infectious diseases and outbreaks: implications for women’s reproductive health and rights in resource-poor settings. Reproductive health, 17(1), 43. 10.1186/s12978-020-0899-y

Cheng, I.-H., Advocat, J., Vasi, S., Enticott, J. C., Willey, S., Wahidi, S., Crock, B., Raghavan, A., Vandenberg, B. E., & Gunatillaka, N. (2018). Report to the World Health Organization April 2018. *World Health*.

Coatsworth, J. D., Pantín, H., McBride, C. K., Briones, E., Kurtines, W. M., & Szapocznik, J. (2002). Ecodevelopmental Correlates of Behavior Problemsin Young Hispanic Females. Applied Developmental Science. 10.1207/s1532480xads0603_3

Cohodes, E. M., Kribakaran, S., Odriozola, P., Bakirci, S., McCauley, S., Hodges, H. R., Sisk, L. M., Zacharek, S. J., & Gee, D. G. (2021). Migration-related trauma and mental health among migrant children emigrating from Mexico and Central America to the United States: Effects on developmental neurobiology and implications for policy. Developmental Psychobiology, 63(6), e22158. 10.1002/dev.22158

Corosky, G. J., & Blystad, A. (2016). Staying Healthy “Under the Sheets”: Inuit Youth Experiences of Access to Sexual and Reproductive Health and Rights in Arviat, Nunavut, Canada. International Journal of Circumpolar Health. 10.3402/ijch.v75.31812

Curtis, E., Jones, R., Tipene[Leach, D., Walker, C., Loring, B., Paine, S. J., & Reid, P. (2019). Why Cultural Safety Rather Than Cultural Competency Is Required to Achieve Health Equity: A Literature Review and Recommended Definition. International journal for equity in health. 10.1186/s12939-019-1082-3

Davis, A. M., Rubinstein, T., Rodriguez, M., & Knight, A. (2017). Mental Health Care for Youth With Rheumatologic Diseases – Bridging the Gap. Pediatric Rheumatology. 10.1186/s12969-017-0214-9

Dey, P., & Sitharthan, G. (2017). Acculturation of Indian Subcontinental Adolescents Living in Australia. Australian Psychologist, 52(3), 238–247. 10.1111/ap.12190

Dulin, A. J., Dale, S. K., Earnshaw, V. A., Fava, J. L., Mugavero, M. J., Napravnik, S., Hogan, J. W., Carey, M. P., & Howe, C. J. (2018). Resilience and HIV: a review of the definition and study of resilience. AIDS Care, 30(sup5), S6–S17. 10.1080/09540121.2018.1515470

Dune, T., Perz, J., Mengesha, Z., & Ayika, D. (2017). Culture Clash? Investigating constructions of sexual and reproductive health from the perspective of 1.5 generation migrants in Australia using Q methodology. Reproductive health, *14*(1), 1-13.

Elkington, K. S., Bauermeister, J. A., Brackis-Cott, E., Dolezal, C., & Mellins, C. A. (2009). Substance Use and Sexual Risk Behaviors in Perinatally Human Immunodeficiency Virus-Exposed Youth: Roles of Caregivers, Peers and HIV Status. Journal of Adolescent Health. 10.1016/j.jadohealth.2009.01.004

Endler, M., Al-Haidari, T., Benedetto, C., Chowdhury, S., Christilaw, J., El Kak, F., Galimberti, D., Garcia-Moreno, C., Gutierrez, M., Ibrahim, S., Kumari, S., McNicholas, C., Mostajo Flores, D., Muganda, J., Ramirez-Negrin, A., Senanayake, H., Sohail, R., Temmerman, M., & Gemzell-Danielsson, K. (2021). How the coronavirus disease 2019 pandemic is impacting sexual and reproductive health and rights and response: Results from a global survey of providers, researchers, and policy-makers. Acta Obstetricia et Gynecologica Scandinavica, 100(4), 571–578. 10.1111/aogs.14043

Fair, F., Soltani, H., Raben, L., Streun, Y. v., Sioti, E., Papadakaki, M., Burke, C., Watson, H., Jokinen, M., Shaw, E., Triantafyllou, E., Muijsenbergh, M. v. d., & Vivilaki, V. (2021). Midwives’ Experiences of Cultural Competency Training and Providing Perinatal Care for Migrant Women a Mixed Methods Study: Operational Refugee and Migrant Maternal Approach (ORAMMA) Project. BMC Pregnancy and Childbirth. 10.1186/s12884-021-03799-1

Family Planning, V. (2016). Annual Report 2015-16. W. Print. https://shvic.org.au/assets/resources/FPV_Annual_Report_21_DIGITAL.pdf

Fernandes, B., Cherrett, C., Moryosef, L., Lau, N., & Wykes, J. (2017). Nasopharyngeal Carcinoma: Knowledge Amongst General Practitioners in Western Sydney. Journal of Community Medicine & Health Education. 10.4172/2161-0711.1000517

Ganczak, M., Czubińska, G., Korzeń, M., & Szych, Z. (2017). A Cross-Sectional Study on Selected Correlates of High risk Sexual Behavior in Polish Migrants Resident in the United Kingdom. International journal of environmental research and public health, 14(4), 422. https://www.mdpi.com/1660-4601/14/4/422

Gazard, B., Chui, Z., Harber-Aschan, L., MacCrimmon, S., Bakolis, I., Rimes, K. A., Hotopf, M., & Hatch, S. L. (2018). Barrier or Stressor? The Role of Discrimination Experiences in Health Service Use. BMC public health. 10.1186/s12889-018-6267-y

Gifford, W., Dick, P., Larocque, C., Modanloo, S., Wazni, L., Awar, Z. A., & Benoit, M. (2023). What Culturally Safe Cancer Care Means to Algonquins of Pikwakanagan First Nation. Alternative an International Journal of Indigenous Peoples. 10.1177/11771801231168681

Gifford, W., Thomas, R., Barton, G., & Graham, I. D. (2019). Providing Culturally Safe Cancer Survivorship Care With Indigenous Communities: Study Protocol for an Integrated Knowledge Translation Study. Pilot and Feasibility Studies. 10.1186/s40814-019-0422-9

Gonzales, G., Przedworski, J., & Henning-Smith, C. (2016). Comparison of Health and Health Risk Factors Between Lesbian, Gay, and Bisexual Adults and Heterosexual Adults in the United States: Results From the National Health Interview Survey. JAMA Internal Medicine, 176(9), 1344–1351. 10.1001/jamainternmed.2016.3432

Goodwin, L., Gazard, B., Aschan, L., MacCrimmon, S., Hotopf, M., & Hatch, S. L. (2017). Taking an Intersectional Approach to Define Latent Classes of Socioeconomic Status, Ethnicity and Migration Status for Psychiatric Epidemiological Research. Epidemiology and Psychiatric Sciences. 10.1017/s2045796017000142

Gray, C., Crawford, G., Maycock, B., & Lobo, R. (2021). Socioecological Factors Influencing Sexual Health Experiences and Health Outcomes of Migrant Asian Women Living in ‘Western’ High-Income Countries: A Systematic Review. International journal of environmental research and public health. 10.3390/ijerph18052469

Haas, B. d., Hutter, I., & Timmerman, G. (2017). Young People’s Perceptions of Relationships and Sexual Practices in the Abstinence-Only Context of Uganda. Sex Education. 10.1080/14681811.2017.1315933

Habtamu, D., & Adamu, A. (2013). Assessment of Sexual and Reproductive Health Status of Street Children in Addis Ababa. Journal of Sexually Transmitted Diseases, 2013, 524076. 10.1155/2013/524076

Hameed, S., Maddams, A., Lowe, H., Davies, L., Khosla, R., & Shakespeare, T. (2020). From Words to Actions: Systematic Review of Interventions to Promote Sexual and Reproductive Health of Persons With Disabilities in Low- And Middle-Income Countries. BMJ Global Health. 10.1136/bmjgh-2020-002903

Hawkey, A., Ussher, J. M., & Perz, J. (2021). What Do Women Want? Migrant and Refugee Women’s Preferences for the Delivery of Sexual and Reproductive Healthcare and Information. Ethnicity and Health. 10.1080/13557858.2021.1980772

Haynes, V., Calgaro, E., & Dominey-Howes, D. (2021). The Future of Our Suburbs: Analyses of Heatwave Vulnerability in a Planned Estate. Geographical Research. 10.1111/1745-5871.12498

Herd, P., Higgins, J., Sicinski, K., & Merkurieva, I. (2016). The Implications of Unintended Pregnancies for Mental Health in Later Life. American Journal of Public Health, 106(3), 421–429. 10.2105/ajph.2015.302973

Herrick, A. L., Stall, R., Goldhammer, H., Egan, J. E., & Mayer, K. H. (2013). Resilience as a Research Framework and as a Cornerstone of Prevention Research for Gay and Bisexual Men: Theory and Evidence. Aids and Behavior. 10.1007/s10461-012-0384-x

Heslehurst, N., Brown, H., Pemu, A., Coleman, H., & Rankin, J. (2018). Perinatal Health Outcomes and Care Among Asylum Seekers and Refugees: A Systematic Review of Systematic Reviews. BMC Medicine. 10.1186/s12916-018-1064-0

Horyniak, D., Melo, J. S., Farrell, R. M., Ojeda, V. D., & Strathdee, S. A. (2016). Epidemiology of Substance Use Among Forced Migrants: A Global Systematic Review. PLoS One. 10.1371/journal.pone.0159134

Huang, M.-F., Chang, Y. P., Lin, C. Y., & Yen, C. F. (2022). A Newly Developed Scale for Assessing Experienced and Anticipated Sexual Stigma in Health-Care Services for Gay and Bisexual Men. International journal of environmental research and public health. 10.3390/ijerph192113877

Iqbal, S., Zakar, R., Zakar, M. Z., & Fischer, F. (2017). Perceptions of Adolescents’ Sexual and Reproductive Health and Rights: A Cross-Sectional Study in Lahore District, Pakistan. BMC International Health and Human Rights. 10.1186/s12914-017-0113-7

Ivanova, O., Rai, M., & Kemigisha, E. (2018). A Systematic Review of Sexual and Reproductive Health Knowledge, Experiences and Access to Services among Refugee, Migrant and Displaced Girls and Young Women in Africa. International journal of environmental research and public health, 15(8), 1583. https://www.mdpi.com/1660-4601/15/8/1583

James, P. B., Wardle, J., Steel, A., & Adams, J. (2020). An assessment of Ebola-related stigma and its association with informal healthcare utilisation among Ebola survivors in Sierra Leone: a cross-sectional study. BMC public health, 20(1), 182. 10.1186/s12889-020-8279-7

Josefsson, K. A., Schindele, A. C., Deogan, C., & Lindroth, M. (2019). Education for Sexual and Reproductive Health and Rights (SRHR): A Mapping of SRHR-related Content in Higher Education in Health Care, Police, Law and Social Work in Sweden. Sex Education. 10.1080/14681811.2019.1572501

Kagitcibasi, C. (2007). Family, self, and human development across cultures: Theory and applications. Routledge.

Kalra, G., & Bhugra, D. (2013). Sexual Violence Against Women: Understanding Cross-Cultural Intersections. Indian Journal of Psychiatry. 10.4103/0019-5545.117139

Karatay, G., Bowers, B. J., Karadağ, E., & Demir, M. (2016). Cultural Perceptions and Clinical Experiences of Nursing Students in Eastern Turkey. International Nursing Review. 10.1111/inr.12321

Keygnaert, I., & Guieu, A. (2015). What the eye does not see: a critical interpretive synthesis of European Union policies addressing sexual violence in vulnerable migrants. Reproductive Health Matters, 23(46), 45–55.

Khan, M. D., Daniyal, M., Abid, K., Tawiah, K., Tebha, S. S., & Essar, M. Y. (2022). Analysis of Adolescents’ Perception and Awareness Level for Sexual and Reproductive Health Rights in Pakistan. *Health Science Reports*. 10.1002/hsr2.982

Kwankye, S. O., & Augustt, E. (2013). Media Exposure and Reproductive Health Behaviour Among Young Females in Ghana. African Population Studies. 10.11564/22-2-330

Li, S., Huang, H., Xu, G., Cai, Y., Huang, F., & Ye, X. (2013). Substance Use, Risky Sexual Behaviors, and Their Associations in a Chinese Sample of Senior High School Students. BMC public health. 10.1186/1471-2458-13-295

Liamputtong, P. (2006). Researching the vulnerable: A guide to sensitive research methods. Researching the Vulnerable, 1-256.

Liamputtong, P. (2020). Qualitative research methods.

Logie, C. H., Okumu, M., Mwima, S., Kyambadde, P., Hakiza, R., Kibathi, I. P., Kironde, E., Musinguzi, J., & Kipenda, C. U. (2019). Exploring Associations Between Adolescent Sexual and Reproductive Health Stigma and HIV Testing Awareness and Uptake Among Urban Refugee and Displaced Youth in Kampala, Uganda. Sexual and Reproductive Health Matters. 10.1080/26410397.2019.1695380

Lonne, B., Flemington, T., Lock, M., Hartz, D., Ramanathan, S., & Fraser, J. (2020). The Power of Authenticity and Cultural Safety at the Intersection of Healthcare and Child Protection. International Journal on Child Maltreatment Research Policy and Practice. 10.1007/s42448-020-00053-7

Lynch, L., Moorhead, A., Long, M., & Hawthorne-Steele, I. (2021). What Type of Helping Relationship Do Young People Need? Engaging and Maintaining Young People in Mental Health Care—A Narrative Review. Youth & Society, 53(8), 1376–1399. 10.1177/0044118x20902786

Maheen, H., Chalmers, K., Khaw, S., & McMichael, C. (2021). Sexual and Reproductive Health Service Utilisation of Adolescents and Young People From Migrant and Refugee Backgrounds in High-Income Settings: A Qualitative Evidence Synthesis (QES). Sexual health. 10.1071/sh20112

Malia, M., Goleen, S., Jennifer, O., Clarisa, B., & Terry, M. (2021). ‘Scrambling to figure out what to do’: a mixed method analysis of COVID-19’s impact on sexual and reproductive health and rights in the United States. BMJ Sexual & Reproductive Health, 47(4), e16. 10.1136/bmjsrh-2021-201081

Mbarushimana, V., Conco, D. N., & Goldstein, S. (2022). “Such conversations are not had in the families”: a qualitative study of the determinants of young adolescents’ access to sexual and reproductive health and rights information in Rwanda. BMC public health, 22(1), 1–14.

McCann, T. V., Mugavin, J., Renzaho, A. Μ. Ν., & Lubman, D. I. (2016). Sub-Saharan African Migrant Youths’ Help-Seeking Barriers and Facilitators for Mental Health and Substance Use Problems: A Qualitative Study. BMC Psychiatry. 10.1186/s12888-016-0984-5

Mengesha, Z. B., Perz, J., Dune, T., & Ussher, J. (2017). Refugee and migrant women’s engagement with sexual and reproductive health care in Australia: A socio-ecological analysis of health care professional perspectives. PLoS One, 12(7), e0181421. 10.1371/journal.pone.0181421

Mengesha, Z. B., Perz, J., Dune, T., & Ussher, J. (2018). Challenges in the provision of sexual and reproductive health care to refugee and migrant women: AQ methodological study of health professional perspectives. Journal of Immigrant and Minority Health, 20(2), 307–316.

Mengesha, Z. B., Perz, J., Dune, T., & Ussher, J. M. (2018). Preparedness of Health Care Professionals for Delivering Sexual and Reproductive Health Care to Refugee and Migrant Women: A Mixed Methods Study. *International journal of environmental research and public health*. 10.3390/ijerph15010174

Metusela, C., Ussher, J. M., Perz, J., Hawkey, A., Morrow, M. R., Narchal, R., Estoesta, J., & Monteiro, M. (2017). “In My Culture, We Don’t Know Anything About That”: Sexual and Reproductive Health of Migrant and Refugee Women. International journal of behavioral medicine. 10.1007/s12529-017-9662-3

Miller, D. J., McBain, K. A., Li, W. W., & Raggatt, P. T. F. (2019). Pornography, preference for porn-like sex, masturbation, and men’s sexual and relationship satisfaction. Personal Relationships, 26(1), 93–113. 10.1111/pere.12267

Mittal, M., Senn, T. E., & Carey, M. P. (2013). Fear of Violent Consequences and Condom Use Among Women Attending an STD Clinic. Women & Health. 10.1080/03630242.2013.847890

Mmari, K., Lantos, H., Brahmbhatt, H., Delany-Moretlwe, S., Lou, C., Acharya, R., & Sangowawa, A. (2014). How adolescents perceive their communities: a qualitative study that explores the relationship between health and the physical environment. BMC public health, 14(1), 1–12.

Mpofu, E. (2018). How Religion Frames Health Norms: A Structural Theory Approach: How Religion Frames Health Norms: A Structural Theory Approach. Religions, 9(4), 119.

Mpofu, E., Nkomazana, F., Muchado, J. A., Togarasei, L., & Bingenheimer, J. B. (2014). Faith and HIV prevention: the conceptual framing of HIV prevention among Pentecostal Batswana teenagers. BMC public health, 14(1), 1–11.

Mulubwa, C., Hurtig, A.-K., Zulu, J. M., Michelo, C., Sandøy, I. F., & Goicolea, I. (2020). Can sexual health interventions make community-based health systems more responsive to adolescents? A realist informed study in rural Zambia. Reproductive health, 17(1), 1. 10.1186/s12978-019-0847-x

Napier-Raman, S., Hossain, S. Z., Lee, M.-J., Mpofu, E., Liamputtong, P., & Dune, T. (2023). Migrant and refugee youth perspectives on sexual and reproductive health and rights in Australia: a systematic review. Sexual health, 20(1), 35–48. 10.1071/SH22081

Neal, J. W., & Neal, Z. P. (2013). Nested or Networked? Future Directions for Ecological Systems Theory. Social Development, 22(4), 722–737. 10.1111/sode.12018

Ogbe, E., Braeckel, D. V., Temmerman, M., Larsson, E. C., Keygnaert, I., Aragón, W. d. l. R., Cheng, F., Lazdāne, G., Cooper, D., Shamu, S., Gichangi, P., Dias, S., Barrett, H., Nobels, A., Pei, K., Galle, A., Esho, T., Knight, L., Tabana, H., & Degomme, O. (2018). Opportunities for Linking Research to Policy: Lessons Learned From Implementation Research in Sexual and Reproductive Health Within the ANSER network. Health Research Policy and Systems. 10.1186/s12961-018-0397-7

Phinney, J. S. (1990). Ethnic identity in adolescents and adults: review of research. Psychological bulletin, 108(3), 499.

Pound, P., Langford, R., & Campbell, R. (2016). What Do Young People Think About Their School-Based Sex and Relationship Education? A Qualitative Synthesis of Young People’s Views and Experiences. BMJ Open. 10.1136/bmjopen-2016-011329

Prather, C., Fuller, T. R., Marshall, K. J., & Jeffries, W. L. (2016). The Impact of Racism on the Sexual and Reproductive Health of African American Women. Journal of Women S Health. 10.1089/jwh.2015.5637

Quirkos, L. (2021). Quirkos. In: Quirkos Software Edinburgh.

Rawson, H. A., & Liamputtong, P. (2010). Culture and sex education: the acquisition of sexual knowledge for a group of Vietnamese Australian young women. Ethnicity & Health, 15(4), 343–364. 10.1080/13557851003728264

Reason, P., & Bradbury, H. (2008). The SAGE handbook of action research.

Riza, E., Karnaki, P., Gil-Salmerón, A., Zota, K., Ho, M., Petropoulou, M., Katsas, K., Garcés-Ferrer, J., & Linos, A. (2020). Determinants of Refugee and Migrant Health Status in 10 European Countries: The Mig-HealthCare Project. International journal of environmental research and public health, 17(17), 6353. https://www.mdpi.com/1660-4601/17/17/6353

Roberts, M., Lobo, R., & Sorenson, A. (2017). Evaluating the Sharing Stories youth theatre program: an interactive theatre and drama-based strategy for sexual health promotion among multicultural youth. Health Promotion Journal of Australia, 28(1), 30–36. 10.1071/HE15096

Roseby, R., Adams, K., Leech, M., Taylor, K., & Campbell, D. (2019). Not just a policy; this is for real. An affirmative action policy to encourage Aboriginal and Torres Strait Islander peoples to seek employment in the health workforce. Internal Medicine Journal, 49(7), 908–910. 10.1111/imj.14345

Ruane-McAteer, E., Amin, A., Hanratty, J., Lynn, F., Willenswaard, K. C. v., Reid, E., Khosla, R., & Lohan, M. (2019). Interventions Addressing Men, Masculinities and Gender Equality in Sexual and Reproductive Health and Rights: An Evidence and Gap Map and Systematic Review of Reviews. BMJ Global Health. 10.1136/bmjgh-2019-001634

Saleem, H., Narasimhan, M., Denison, J. A., & Kennedy, C. E. (2017). Achieving Pregnancy Safely for HIV[serodiscordant Couples: A Social Ecological Approach. Journal of the International Aids Society. 10.7448/ias.20.2.21331

Schaaf, M., & Khosla, R. (2021). Necessary but Not Sufficient: A Scoping Review of Legal Accountability for Sexual and Reproductive Health in Low-Income and Middle-Income Countries. BMJ Global Health. 10.1136/bmjgh-2021-006033

Scherer, N., Mactaggart, I., Huggett, C., Pheng, P., Rahman, M.-u., Biran, A., & Wilbur, J. (2021). The Inclusion of Rights of People With Disabilities and Women and Girls in Water, Sanitation, and Hygiene Policy Documents and Programs of Bangladesh and Cambodia: Content Analysis Using EquiFrame. International journal of environmental research and public health. 10.3390/ijerph18105087

Schmitt, É., Hinner, J., & Kruse, A. (2015). Potentials of Survivors, Intergenerational Dialogue, Active Ageing and Social Change. Procedia - Social and Behavioral Sciences. 10.1016/j.sbspro.2015.01.082

Sommet, N., & Berent, J. (2022). Porn Use and Men’s and Women’s Sexual Performance: Evidence From a Large Longitudinal Sample. Psychological Medicine. 10.1017/s003329172100516x

Thornicroft, G., Tansella, M., & Law, A. (2008). Steps, challenges and lessons in developing community mental health care. World Psychiatry, 7(2), 87–92. 10.1002/j.2051-5545.2008.tb00161.x

Tirado, V., Chu, J., Hanson, C., Ekström, A. M., & Kågesten, A. (2020). Barriers and facilitators for the sexual and reproductive health and rights of young people in refugee contexts globally: A scoping review. PLoS One, 15(7), e0236316. 10.1371/journal.pone.0236316

Tirado, V., Engberg, S., Holmblad, I. S., Strömdahl, S., Ekström, A. M., & Hurtig, A. K. (2022). “One-time interventions, it doesn’t lead to much” – healthcare provider views to improving sexual and reproductive health services for young migrants in Sweden. BMC Health Services Research, 22(1), 668. 10.1186/s12913-022-07945-z

Torke, A. M., & Carnahan, J. L. (2017). Optimizing the Clinical Care of Lesbian, Gay, Bisexual, and Transgender Older Adults. JAMA Internal Medicine. 10.1001/jamainternmed.2017.5324

Ussher, J. M., Perz, J., Metusela, C., Hawkey, A. J., Morrow, M., Narchal, R., & Estoesta, J. (2017). Negotiating discourses of shame, secrecy, and silence: Migrant and refugee women’s experiences of sexual embodiment. Archives of Sexual Behavior, 46(7), 1901–1921.

VanderWielen, L. M., Enurah, A. S., Rho, H. Y., Nagarkatti-Gude, D. R., Michelsen-King, P., Crossman, S. H., & Vanderbilt, A. A. (2014). Medical Interpreters. Academic Medicine. 10.1097/acm.0000000000000296

Veenstra, G. (2011). Race, Gender, Class, and Sexual Orientation: Intersecting Axes of Inequality and Self-Rated Health in Canada. International journal for equity in health. 10.1186/1475-9276-10-3

Villa-Torres, L., & Svanemyr, J. (2015). Ensuring Youth’s Right to Participation and Promotion of Youth Leadership in the Development of Sexual and Reproductive Health Policies and Programs. Journal of Adolescent Health. 10.1016/j.jadohealth.2014.07.022

Wado, Y. D., Bangha, M., Kabiru, C. W., & Feyissa, G. T. (2020). Nature Of, and Responses to Key Sexual and Reproductive Health Challenges for Adolescents in Urban Slums in Sub-Saharan Africa: A Scoping Review. Reproductive health. 10.1186/s12978-020-00998-5

Wright, P. J., Paul, B., Herbenick, D., & Tokunaga, R. S. (2021). Pornography and Sexual Dissatisfaction: The Role of Pornographic Arousal, Upward Pornographic Comparisons, and Preference for Pornographic Masturbation. Human Communication Research. 10.1093/hcr/hqab001

Yakushko, O., Watson, M., & Thompson, S. (2008). Stress and Coping in the Lives of Recent Immigrants and Refugees: Considerations for Counseling. International Journal for the Advancement of Counselling, 30(3), 167–178. 10.1007/s10447-008-9054-0

Zulu, J. M., Goicolea, I., Kinsman, J., Sandøy, I. F., Blystad, A., Mulubwa, C., Makasa, M., Michelo, C., Musonda, P., & Hurtig, A.-K. (2018). Community Based Interventions for Strengthening Adolescent Sexual Reproductive Health and Rights: How Can They Be Integrated and Sustained? A Realist Evaluation Protocol From Zambia. Reproductive health. 10.1186/s12978-018-0590-8

